# Resource allocation strategies for insecticide-treated bed nets to achieve malaria eradication

**DOI:** 10.1101/2023.04.16.23288647

**Authors:** Nora Schmit, Hillary M Topazian, Matteo Pianella, Giovanni D Charles, Peter Winskill, Michael T White, Katharina Hauck, Azra C Ghani

**Author notes:** These authors contributed equally to this work (Nora Schmit, Hillary M Topazian, Matteo Pianella). <u>Corresponding author</u>: Nora Schmit, MRC Centre for Global Infectious Disease Analysis Imperial College London, St Mary’s Campus, Norfolk Place, London W2 1PG.

## Abstract

**Background:** Large reductions in the global malaria burden have been achieved in the last decades, but plateauing funding poses a challenge for progressing towards the ultimate goal of malaria eradication. We aimed to determine the optimal strategy to allocate global resources to achieve this goal.

**Methods:** Using previously published mathematical models of *Plasmodium falciparum* and *Plasmodium vivax* transmission incorporating insecticide-treated nets (ITNs) as an illustrative intervention, we sought to identify the global funding allocation that maximized impact under defined objectives and across a range of global funding budgets.

**Results:** We found that the optimal strategy for case reduction closely mirrored an allocation framework that prioritizes funding for high-transmission settings, resulting in total case reductions of 76% (optimal strategy) and 66% (prioritizing high-transmission settings) at intermediate budget levels. Allocation strategies that had the greatest impact on case reductions were associated with lesser near-term impacts on the global population at risk, highlighting a trade-off between reducing burden and “shrinking the map” through a focus on near-elimination settings. The optimal funding distribution prioritized high ITN coverage in high-transmission settings endemic for *P. falciparum* only, while maintaining lower levels in low-transmission settings. However, at high budgets, 62% of funding was targeted to low-transmission settings co-endemic for *P. falciparum* and *P. vivax*.

**Conclusions:** These results support current global strategies to prioritize funding to high-burden *P. falciparum*-endemic settings in sub-Saharan Africa to minimize clinical malaria burden and progress towards elimination but highlight competing goals of reducing the global population at risk and addressing the burden of *P. vivax*.

## INTRODUCTION

Global support for malaria eradication has fluctuated in response to changing health policies over the past 75 years. From near global endemicity in the 1900’s over 100 countries have eliminated malaria, with 10 of these certified malaria-free by the World Health Organization (WHO) in the last two decades (Feachem et al., 2010, Shretta et al., 2017, Weiss et al., 2019). Despite this success, 41% and 57% of the global population in 2017 were estimated to live in areas at risk of infection with *Plasmodium falciparum* and *Plasmodium vivax*, respectively (Weiss et al., 2019, Battle et al., 2019). In 2021 there were an estimated 247 million new malaria cases and over 600,000 deaths, primarily in children under 5 years of age (World Health Organization, 2022b). Mosquito resistance to the insecticides used in vector control, parasite resistance to both first-line therapeutics and diagnostics, and local active conflicts continue to threaten elimination efforts (World Health Organization, 2020). Nevertheless, the global community continues to strive towards the ultimate aim of eradication, which could save millions of lives and thus offer high returns on investment (Chen et al., 2018, Strategic Advisory Group on Malaria Eradication, 2020).

The global goals outlined in the Global Technical Strategy for Malaria (GTS) 2016-2030 include reducing malaria incidence and mortality rates by 90%, achieving elimination in 35 countries, and preventing re-establishment of transmission in all countries currently classified as malaria-free by 2030 (World Health Organization, 2015). Various stakeholders have also set timelines for the wider goal of global eradication, ranging from 2030 to 2050 (World Health Organization, 2020, Chen et al., 2018, Strategic Advisory Group on Malaria Eradication, 2020). However, there remains a lack of consensus on how best to achieve this longer-term aspiration. Historically, large progress was made in eliminating malaria mainly in lower-transmission countries in temperate regions during the Global Malaria Eradication Program in the 1950s, with the global population at risk of malaria reducing from around 70% of the world population in 1950 to 50% in 2000 (Hay et al., 2004). Renewed commitment to malaria control in the early 2000s with the Roll Back Malaria initiative subsequently extended the focus to the highly endemic areas in sub-Saharan Africa (Feachem et al., 2010). Whilst it is now widely acknowledged that the current tool set is insufficient in itself to eradicate the parasite, there continues to be debate about how resources should be allocated (Snow, 2015). Some advocate for a focus on high-burden settings to lower the overall global burden (World Health Organization, 2019), while others call for increased funding to middle-income low-burden countries through a ‘shrink the map strategy’ where elimination is considered a driver of global progress (Newby et al., 2016). A third set of policy options is influenced by equity considerations including allocating funds to achieve equal allocation per person at risk, equal access to bed nets and treatment, maximize lives saved, or to achieve equitable overall health status (World Health Organization, 2013, Raine et al., 2016).

Global strategies are influenced by international donors, which represent 68% of the global investment in malaria control and elimination activities (World Health Organization, 2022b). The Global Fund and the U.S. President’s Malaria Initiative are two of the largest contributors to this investment. Their strategies pursue a combination approach, prioritizing malaria reduction in high-burden countries while achieving sub-regional elimination in select settings (The Global Fund, 2021, United States Agency for International Development and Centers for Disease Control and Prevention, 2021). Given that the global investment for malaria control and elimination still falls short of the 6.8 billion USD currently estimated to be needed to meet GTS 2016-2030 goals (World Health Organization, 2020), an optimized strategy to allocate limited resources is critical to maximizing the chance of successfully achieving the GTS goals and longer-term eradication aspirations.

In this study, we use mathematical modelling to explore the optimal allocation of limited global resources to maximize the long-term reduction in *P. falciparum* and *P. vivax* malaria. Our aim is to determine whether financial resources should initially focus on high-transmission countries, low-transmission countries, or a balance between the two across a range of global budgets. In doing so, we consider potential trade-offs between short-term gains and long-term impact. We use compartmental deterministic versions of two previously developed and tested individual-based model of *P. falciparum* and *P. vivax* transmission respectively (Griffin et al., 2010, White et al., 2018). Using the compartmental model structures allows us to fully explore the space of possible resource allocation decision using optimization, which would be prohibitively costly to perform using more complex individual-based models. Furthermore, to evaluate the impact of resource allocation options, we focus on a single intervention - insecticide-treated nets (ITNs). Whilst in reality national malaria elimination programs encompass a broad range of preventative and therapeutic tools alongside different surveillance strategies as transmission decreases, this simplification is made for computational feasibility, with ITNs chosen as they (a) provide both an individual protective effect and population-level transmission reductions (i.e. indirect effects); (b) are the most widely used single malaria intervention other than first-line treatment; and (c) extensive distribution and costing data are available that allow us to incorporate their decreasing technical efficiency at high coverage.

## RESULTS

We identified 105 malaria-endemic countries based on 2000 *P. falciparum* and *P. vivax* prevalence estimates (before the scale-up of interventions), of which 44, 9, and 52 were endemic for *P. falciparum* only, *P. vivax* only, and co-endemic for both species, respectively. Globally, the clinical burden of malaria was focused in settings of high transmission intensity endemic for *P. falciparum* only, followed by low-transmission settings co-endemic for *P. falciparum* and *P. vivax* (**Figure 1A**). Conversely, 89% of the global population at risk of malaria was located in co-endemic settings with very low and low transmission intensities (**Figure 1B**). All 25 countries with high transmission intensity and 11 of 17 countries with moderate transmission intensity were in Africa, while almost half of global cases and population at risk in low-transmission co-endemic settings originated in India.

**Figure 1.**
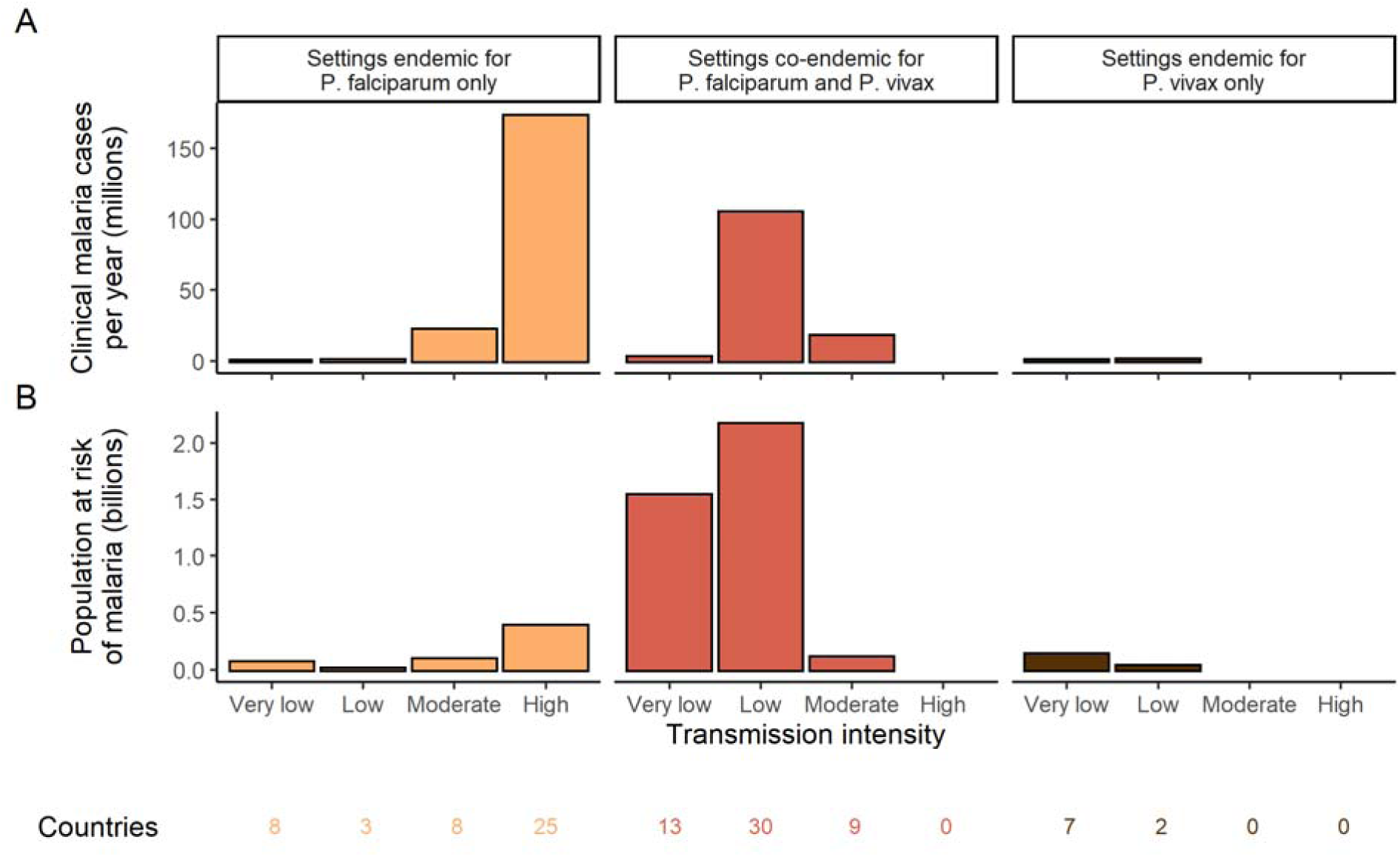
Global distribution of *P. falciparum* and *P. vivax* malaria burden in 2000 (in the absence of insecticide-treated nets) obtained from the Malaria Atlas Project (Weiss et al., 2019, Battle et al., 2019). A) The annual number of clinical cases and B) the population at risk of malaria across settings with different transmission intensities and endemic for *P. falciparum*, *P. vivax* or co-endemic for both species. The number of countries in each setting is indicated below the figure.

Deterministic compartmental versions of two previously published and validated mathematical models of *P. falciparum* and *P. vivax* malaria transmission dynamics (Griffin et al., 2010, Griffin et al., 2014, Griffin et al., 2016, White et al., 2018) were used to explore associations between ITN use and clinical malaria incidence. In model simulations, the relationship between ITN usage and malaria infection outcomes varied by the baseline entomological inoculation rate (EIR), representing local transmission intensity, and parasite species (**Figure 2**). The same increase in ITN usage achieved a larger relative reduction in clinical incidence in low-EIR than in high-EIR settings. Low levels of ITN usage were sufficient to eliminate malaria in low-transmission settings, whereas high ITN usage was necessary to achieve a substantial decrease in clinical incidence in high-EIR settings. At the same EIR value, ITNs also led to a larger relative reduction in *P. falciparum* than *P. vivax* clinical incidence. However, ITN usage of 80% was not sufficient to lead to full elimination of either *P. falciparum* or *P. vivax* in the highest transmission settings. In combination, the models projected that ITNs could reduce global *P. falciparum* and *P. vivax* cases by 83.6% from 252.0 million and by 99.9% from 69.3 million in 2000, respectively, assuming a maximum ITN usage of 80%.

**Figure 2.**
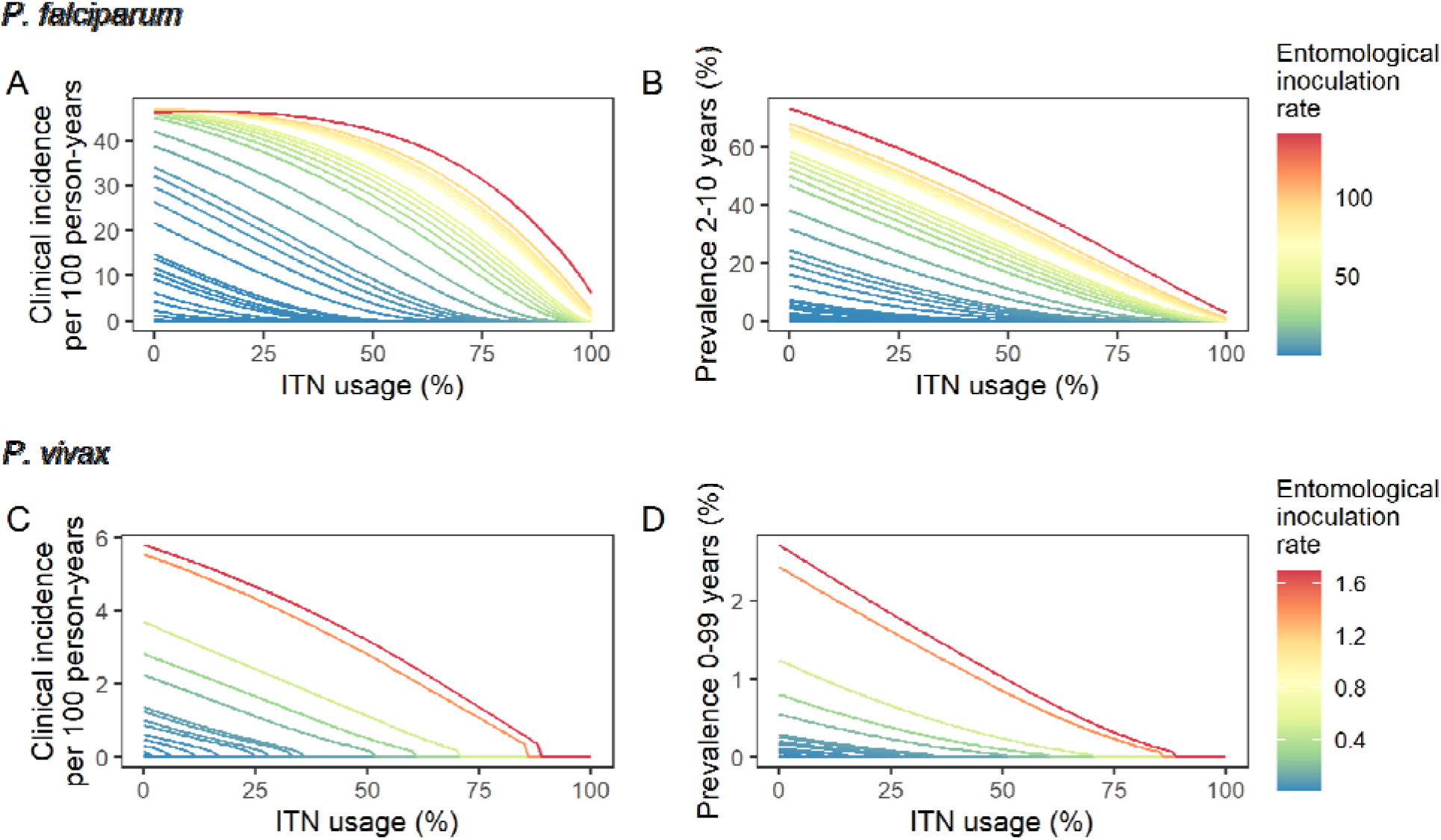
Modelled impact of insecticide-treated net (ITN) usage on malaria epidemiology by the setting-specific transmission intensity, represented by the baseline entomological inoculation rate. The impact on the clinical incidence and prevalence of *P. falciparum* malaria (panels A and B) and on the clinical incidence and prevalence of *P. vivax* malaria (panels C and D) is shown. Panels A and C represent the clinical incidence for all ages.

We next used a non-linear generalized simulated annealing function to determine the optimal global resource allocation for ITNs across a range of budgets. We defined optimality as the funding allocation across countries which minimizes a given objective. We considered two objectives: first, reducing the global number of clinical malaria cases, and second, reducing both the global number of clinical cases and the number of settings not having yet reached a pre-elimination phase. The latter can be interpreted as accounting for an additional positive contribution of progressing towards elimination on top of a reduced case burden (e.g. general health system strengthening through a reduced focus on malaria). To relate funding to impact on malaria, we incorporated a non-linear relationship between costs and ITN usage, resulting in an increase in the marginal cost of ITN distribution at high coverage levels (Bertozzi-Villa et al., 2021). We considered a range of fixed budgets, with the maximum budget being that which enabled achieving the lowest possible number of cases in the model. Low, intermediate and high budget levels refer to 25%, 50% and 75% of this maximum, respectively.

In our main analysis we ignored the time dimension over which funds are distributed, instead focusing on the endemic equilibrium reached for each level of allocation (sensitivity to this assumption is explored in a second analysis with dynamic re-allocation every 3 years). The optimal strategies were compared with three existing approaches to resource allocation: 1) prioritization of high-transmission settings, 2) prioritization of low-transmission (near-elimination) settings, and 3) proportional allocation by disease burden. Strategies prioritizing high- or low-transmission settings involved sequential allocation of funding to groups of countries based on their transmission intensity (from highest to lowest EIR or *vice versa*). The proportional allocation strategy mimics the current allocation algorithm employed by the Global Fund: budget shares are distributed according to malaria disease burden in the 2000-2004 period (The Global Fund, 2019). To allow comparison with this existing funding model, we also started allocation decisions from the year 2000.

We found that the optimal strategies for reducing total malaria cases (i.e. global burden) and for case reduction and pre-elimination to be similar to the strategy that prioritized funding for high-transmission settings. These three strategies achieved the largest reductions in global malaria cases at all budgets, including reductions of 76%, 73% and 66% at the intermediate budget level, respectively (**Figure 3A**, **Table 1**). At low to intermediate budgets, the proportional allocation strategy also reduced malaria cases effectively by up to 53%. While these four scenarios had very similar effects on malaria cases at low budgets, they diverged with increasing funding, where the proportional allocation strategy did not achieve substantial further reductions. Depending on the available budget, the optimal strategy for case reduction averted up to 31% more cases than prioritization of high-transmission settings and 64% more cases than proportional allocation, corresponding to respective differences of 37.9 and 74.5 million cases globally.

**Figure 3.**
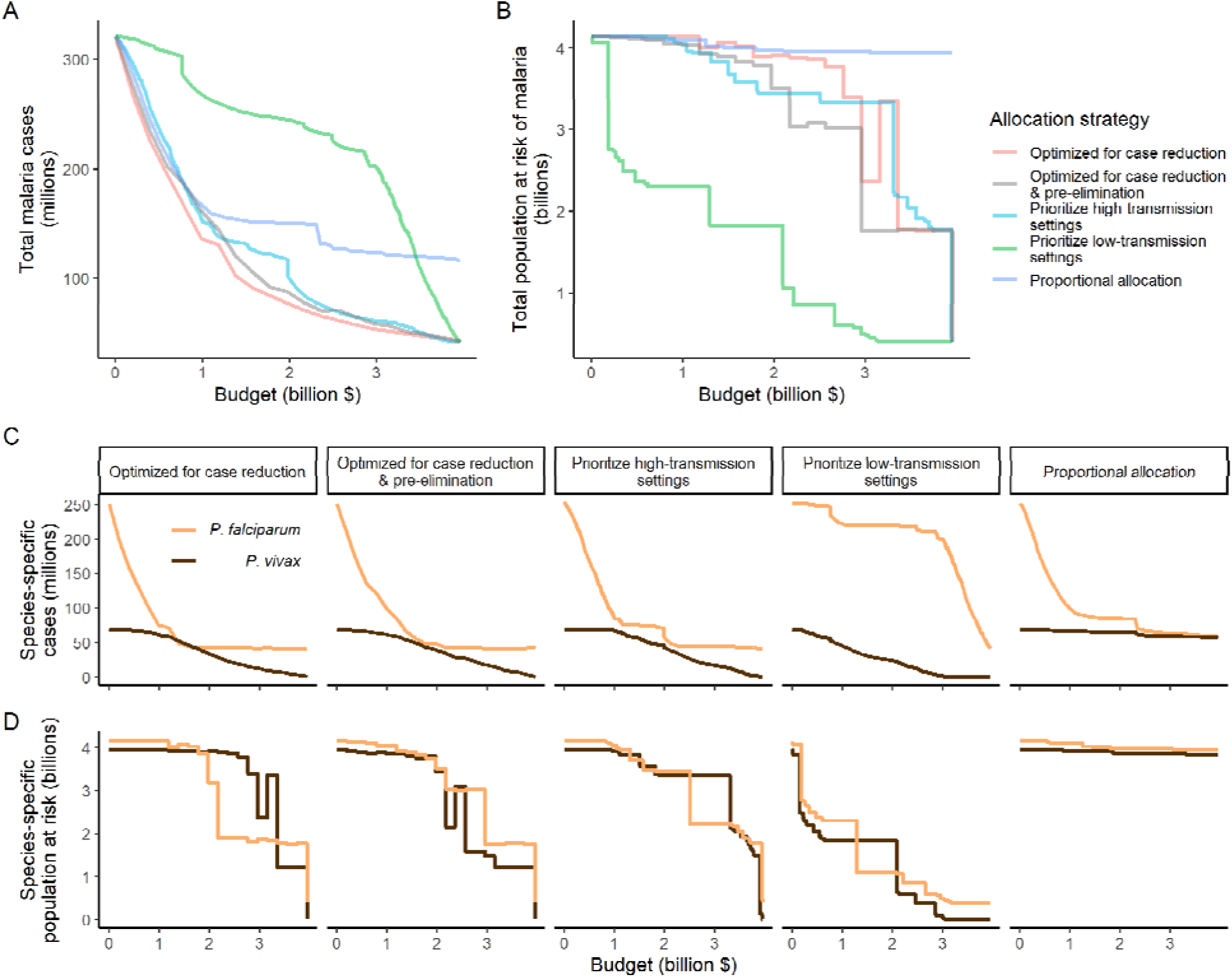
Global clinical cases and population at risk of malaria under different allocation strategies at varying budgets. The impact on total malaria cases (panel A), total population at risk (panel B), individual *P. falciparum* and *P. vivax* cases (panel C) and population at risk of either species (panel D) are shown. Budget levels range from 0, representing no usage of insecticide-treated nets, to the budget required to achieve the maximum possible impact. Optimizing for case reduction generally leads to declining populations at risk as the budget increases, but this is not guaranteed due to the possibility of redistribution of funding between settings to minimize cases. The strategy optimizing case reduction and pre-elimination shown here places the same weighting (1:1) on reaching pre-elimination in a setting as on averting total cases, but conclusions were the same for weights of 0.5-100 on pre-elimination.

**Table 1.**
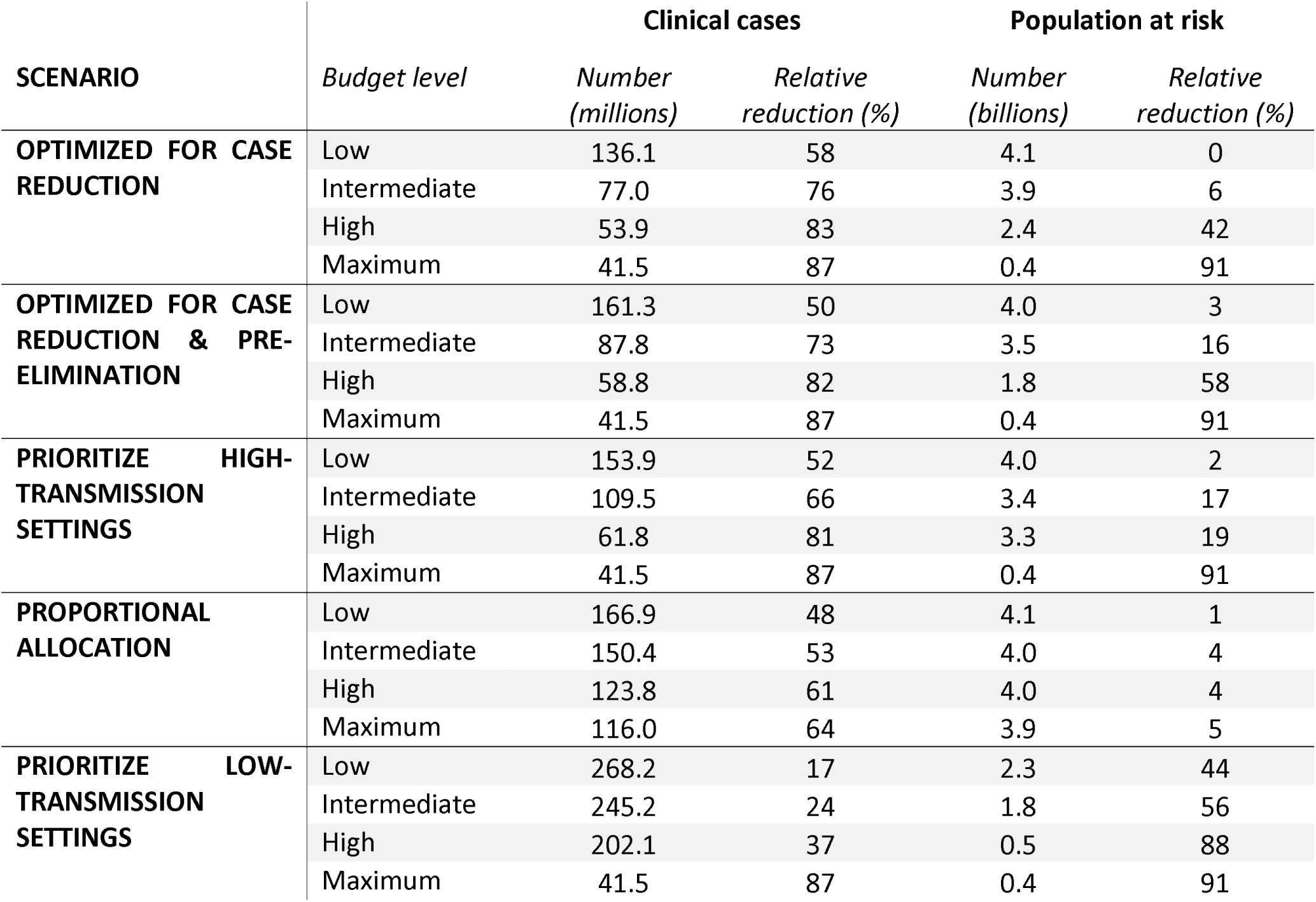
Relative reduction in malaria cases and population at risk under different allocation strategies. Reductions are shown relative to the baseline of 321 million clinical cases and 4.1 billion persons at risk in the absence of interventions. Low, intermediate and high budget levels represent 25%, 50% and 75% of the maximum budget, respectively. The strategy optimizing case reduction and pre-elimination shown here places the same weighting (1:1) on reaching pre-elimination in a setting as on averting total cases.

We additionally found there to be a trade-off between reducing global cases and reducing the global population at risk of malaria. Both the optimal strategies and the strategy prioritizing high-transmission settings did not achieve substantial reductions in the global population at risk until large investments were reached (**Figure 3B**, **Table 1**). Even at a high budget, the global population at risk was only reduced by 19% under the scenario prioritizing high-transmission settings, with higher reductions of 42-58% for the optimal strategies, while proportional allocation had almost no effect on this outcome. Conversely, diverting funding to prioritize low-transmission settings was highly effective at increasing the number of settings eliminating malaria, achieving a 56% reduction in the global population at risk already at intermediate budgets. However, this investment only led to a minimal reduction of 24% in total malaria case load (**Figure 3**, **Table 1**). At high budget levels, prioritizing low-transmission settings resulted in up to 3.8 times (a total of 159.4 million) more cases than the optimal allocation for case reduction. Despite the population at risk remaining relatively large with the optimal strategy for case reduction and pre-elimination, it nevertheless led to pre-elimination in more malaria-endemic settings than all other strategies (**Figure S7**), in addition to close to minimum cases across all budgets (**Figure 3**).

The allocation strategies also had differential impacts on *P. falciparum* and *P. vivax* cases, with case reductions generally occurring first for *P. falciparum* except when prioritizing low-transmission settings. *P. vivax* cases were not substantially affected at low global budgets for all other allocation strategies, and proportional allocation had almost no effect on reducing *P. vivax* clinical burden at any budget (**Figure 3C**), leading to a temporary increase in the proportion of total cases attributable to *P. vivax* relative to *P. falciparum*. The global population at risk remained high with the optimal strategy for case reduction even at high budgets, partly due to a large remaining population at risk of *P. vivax* infection (**Figure 3D**), which was not targeted when aiming to minimize total cases (**Figure 1**).

The optimized distribution of funding to minimize clinical burden depended on the available global budget and was driven by the setting-specific transmission intensity and the population at risk (**Figure 4**, **Figure 1**). With very low to low budget levels, as much as 85% of funding was allocated to moderate to high transmission settings (**Figure 4A, Figure S8A**). This allocation pattern led to the maximum ITN usage of 80% being reached in settings of high transmission intensity and smaller population sizes even at low budgets, while maintaining lower levels in low-transmission settings with larger populations (**Figure 4B, Figure S8B**). The proportion of the budget allocated to low and very low transmission settings increased with increasing budgets, and low transmission settings received the majority of funding at intermediate to maximum budgets. This allocation pattern remained very similar when optimizing for both case reduction and pre-elimination (**Figure S9).** Similar patterns were also observed for the optimized distribution of funding between settings endemic for only *P. falciparum* compared to *P. falciparum* and *P. vivax* co-endemic settings (**Figure 4C-D**), with the former being prioritized at low to intermediate budgets. At the maximum budget, 70% of global funding was targeted at low- and very low-transmission settings co-endemic for both parasite species.

**Figure 4.**
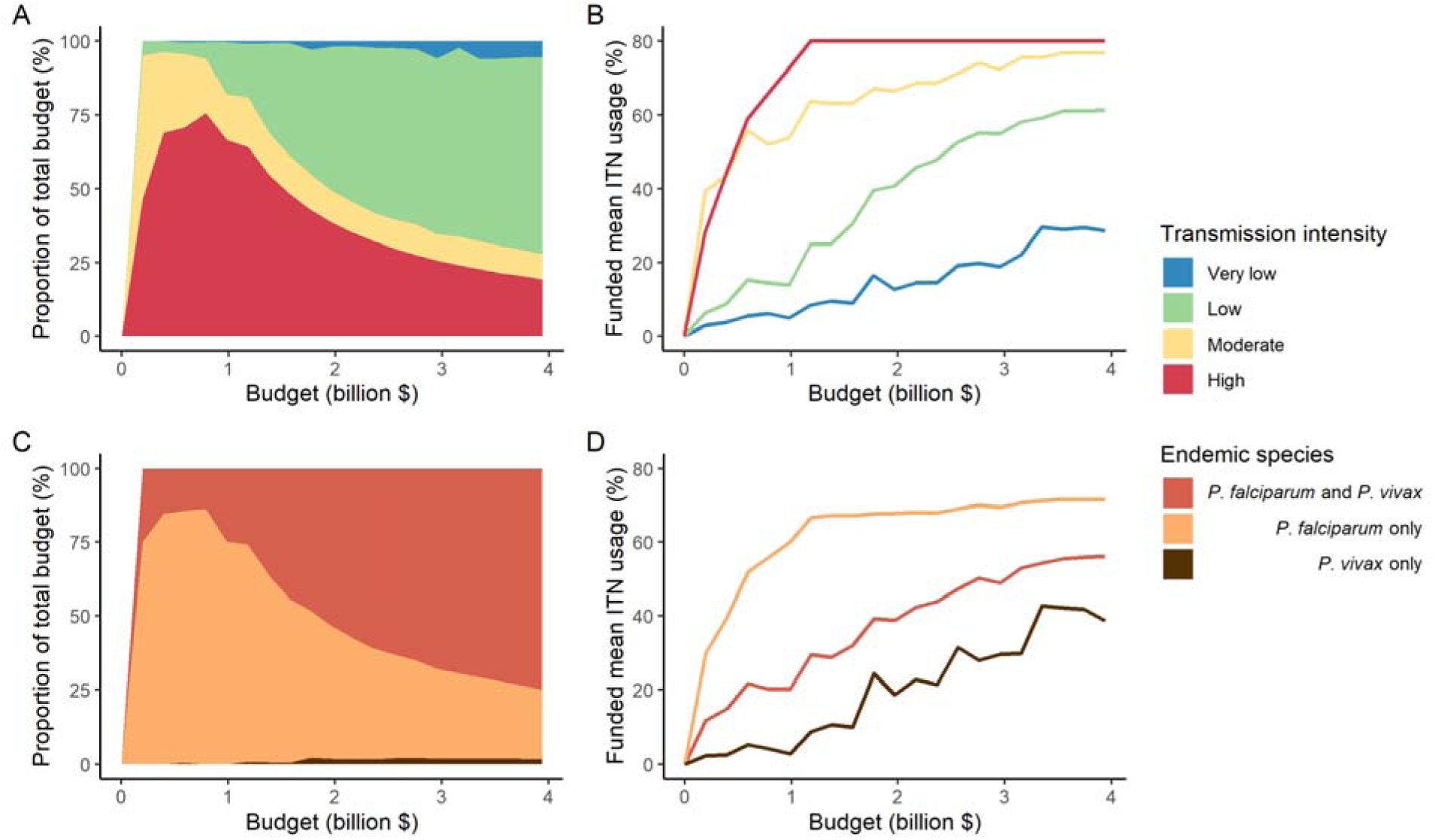
Optimal strategy for funding allocation across settings to minimize malaria case burden at varying budgets. Panels show optimized allocation patterns across settings of different transmission intensity (panels A and B) and different endemic parasite species (panels C and D). The proportion of the total budget allocated to each setting (panels A and C) and the resulting mean population usage of insecticide-treated nets (ITNs) (panels B and D) are shown.

To evaluate the robustness of the results, we conducted a sensitivity analysis on our assumption on ITN distribution efficiency. Results remained similar when assuming a linear relationship between ITN usage and distribution costs (**Figure S10**). While the main analysis involves a single allocation decision to minimise long-term case burden (leading to a constant ITN usage over time in each setting irrespective of subsequent changes in burden), we additionally explored an optimal strategy with dynamic re-allocation of funding every 3 years to minimise cases in the short term. At high budgets, capturing dynamic changes over time through re-allocation of funding based on minimizing *P. falciparum* cases every 3 years led to the same case reductions over time as a one-time optimization with allocation of a constant ITN usage (**Figure S11**). At lower budgets, re-allocation every 3 years achieved a higher impact at several timepoints, but total cases remained similar between the two approaches. Although reallocation of resources from settings which achieved elimination to higher transmission settings did not lead to substantially fewer cases, it reduced total spending over the 39-year period in some cases (**Figure S11**).

## DISCUSSION

Our study highlights the potential impact that funding allocation decisions could have on the global burden of malaria. We estimated that optimizing ITN allocation to minimize global clinical incidence could, at a high budget, avert 83% of clinical cases compared to no intervention. In comparison, the optimal strategy to minimize clinical incidence and maximize the number of settings reaching pre-elimination averted 82% of clinical cases, prioritizing high-transmission settings 81%, proportional allocation 61%, and prioritizing low-transmission settings 37%. Our results support initially prioritizing funding towards reaching high ITN usage in the high-burden *P. falciparum-*endemic settings to minimize global clinical cases and advance elimination in more malaria-endemic settings, but highlight the trade-off between this strategy and reducing the global population at risk of malaria as well as addressing the burden of *P. vivax*.

Prioritizing low-transmission settings demonstrated how focusing on “shrinking the malaria map” by quickly reaching elimination in low-transmission countries diverts funding away from the high-burden countries with the largest caseloads. Prioritizing low-transmission settings achieved elimination in 42% of settings and reduced the global population at risk by 56% when 50% of the maximum budget had been spent, but also resulted in 3.2 times more clinical cases than the optimal allocation scenario. Investing a larger share of global funding towards high-transmission settings aligns more closely with the current WHO “high burden to high impact” approach, which places an emphasis on reducing the malaria burden in the 11 countries which comprise 70% of global cases (World Health Organization, 2019). Previous research supports this approach, finding that the 20 highest burden countries would need to obtain 88% of global investments to reach case and mortality risk estimates in alignment with GTS goals (Patouillard et al., 2017). This is similar to the modelled optimized funding strategy presented here, which allocated up to 76% of very low budgets to settings of high transmission intensity located in sub-Saharan Africa. An initial focus on high- and moderate-transmission settings is further supported by our results showing that a balance can be found between achieving close to optimal case reductions while also progressing towards elimination in the maximum number of settings. Even within a single country, targeting interventions to local hot-spots has been shown to lead to higher cost savings than universal application (Barrenho et al., 2017), and could lead to elimination in settings where untargeted interventions would have little impact (Bousema et al., 2012).

Assessing optimal funding patterns is a global priority due to the funding gap between supply and demand for resources for malaria control and elimination (World Health Organization, 2022b). However, allocation decisions will remain important even if more funding became available, as some of the largest differences in total cases between the modelled strategies occurred at intermediate to high budgets. Our results suggest that most of global funding should only be focused in low-transmission settings co-endemic for *P. falciparum* and *P. vivax* at high budgets once ITN use has already been maximized in high-transmission settings. Global allocation decisions are likely to affect *P. falciparum* and *P. vivax* burden differently, which could have implications for the future global epidemiology of malaria. For example, with a focus on disease burden reduction, a temporary increase in the proportion of malaria cases attributable to *P. vivax* was projected, in line with recent observations in near-elimination areas (Battle et al., 2019, Price et al., 2020). Nevertheless, even when international funding for malaria increased between 2007-2009, African countries remained the major recipients of financial support, while *P. vivax*-dominant countries were not as well funded (Snow et al., 2010). This serves as a reminder that achieving elimination of malaria from all endemic countries will ultimately require targeting investments so as to also address the burden of *P. vivax* malaria.

Different priorities in resource allocation decisions greatly affect which countries receive funding and what health benefits are achieved. The modelled strategies follow key ethical principles in the allocation of scarce healthcare resources, such as targeting those of greatest need (*prioritizing high-transmission settings*, *proportional allocation*) or those with the largest expected health gain (*optimized for case reduction*, *prioritizing high-transmission settings*) (World Health Organization, 2013). Allocation proportional to disease burden did not achieve as great an impact as other strategies because the funding share assigned to settings was constant irrespective of the invested budget and its impact. In modelling this strategy, we did not reassign excess funding in high-transmission settings to other malaria interventions, as would likely occur in practice. This illustrates the possibility that such an allocation approach can potentially target certain countries disproportionally and result in further inequities in health outcomes (Barrenho et al., 2017). From an international funder perspective, achieving vertical equity might therefore also encompass higher disbursements to countries with lower affordability of malaria interventions (Barrenho et al., 2017), as reflected in the Global Fund’s proportional allocation formula which accounts for the economic capacity of countries and specific strategic priorities (The Global Fund, 2019). While these factors were not included in the proportional allocation used here, the estimated impact of these two strategies was nevertheless very similar (**Supplementary Figure S12**).

While our models are based on country patterns of transmission settings and corresponding populations in 2000, there are several factors leading to heterogeneity in transmission dynamics at the national and sub-national level which were not modelled and limit our conclusions. Seasonality, changing population size, and geographic variation in *P. vivax* relapse patterns or in mosquito vectors could affect the projected impact of ITNs and optimized distribution of resources across settings. The two representative *Anopheles* species used in the simulations are also both very anthropophagic, which may have led to overestimation of the effect of ITNs in some settings. By using ITNs as the sole means to reduce mosquito-to-human transmission, we did not capture the complexities of other key interventions that play a role in burden reduction and elimination, the geospatial heterogeneity in cost-effectiveness and optimized distribution of intervention packages on a sub-national level, or related pricing dynamics (Conteh et al., 2021, Drake et al., 2017). For *P. vivax* in particular, reducing the global economic burden and achieving elimination will depend on incorporation of hypnozoitocidal treatment and G6PD screening into case management (Devine et al., 2021). Furthermore, for both parasites, intervention strategies generally become more focal as transmission decreases, with targeted surveillance and response strategies prioritized over widespread vector control. Therefore, policy decisions should additionally be based on analysis of country-specific contexts, and our findings are not informative for individual country allocation decisions. Results do however account for non-linearities in the relationship between ITN distribution and usage to represent changes in cost as a country moves from control to elimination: interventions that are effective in malaria control settings, such as widespread vector control, may be phased out or limited in favor of more expensive active surveillance and a focus on confirmed diagnoses and at-risk populations (Shretta et al., 2017). We also assumed that transmission settings are independent of each other, and did not allow for the possibility of re-introduction of disease, such as has occurred throughout the Eastern Mediterranean from imported cases (World Health Organization Regional Office for the Eastern Mediterranean, 2007). While our analysis presents allocation strategies to progress towards eradication, the results do not provide insight into allocation of funding to maintain elimination. In practice, the threat of malaria resurgence has important implications for when to scale back interventions.

Our analysis demonstrates the most impactful allocation of a global funding portfolio for ITNs to reduce global malaria cases. Unifying all funding sources in a global strategic allocation framework as presented here requires international donor allocation decisions to account for available domestic resources. National governments of endemic countries contribute 31% of all malaria-directed funding globally (World Health Organization, 2020), and government financing is a major source of malaria spending in near-elimination countries in particular (Haakenstad et al., 2019). Within the wider political economy which shapes the funding landscape and priority setting, there remains substantial scope for optimizing allocation decisions, including improving efficiency of within-country allocation of malaria interventions. Subnational malaria elimination in localized settings within a country can also provide motivation for continued elimination in other areas and friendly competition between regions to boost global elimination efforts (Lindblade and Kachur, 2020). Although more efficient allocation cannot fully compensate for projected shortfalls in malaria funding, mathematical modelling can aid efforts in determining optimal approaches to achieve the largest possible impact with available resources.

## MATERIALS AND METHODS

### Transmission models

We used deterministic compartmental versions of two previously published individual-based transmission models of *P. falciparum* and *P. vivax* malaria to estimate the impact of varying ITN usage on clinical incidence in different transmission settings. The *P. falciparum* model has previously been fitted to age-stratified data from a variety of sub-Saharan African settings to recreate observed patterns in parasite prevalence (*PfPR_2-10_*), the incidence of clinical disease, immunity profiles, and vector components relating rainfall, mosquito density, and the entomological inoculation rate (EIR) (Griffin et al., 2016). We developed a deterministic version of an existing individual-based model of *P. vivax* transmission, originally calibrated to data from Papua New Guinea but also shown to reproduce global patterns of *P. vivax* prevalence and clinical incidence (White et al., 2018). Models for both parasite species are structured by age and heterogeneity in exposure to mosquito bites, and account for human immunity patterns. They model mosquito transmission and population dynamics, and the impact of scale-up of ITNs in identical ways. Full assumptions, mathematical details and parameter values can be found in the Supplementary Material and in previous publications (Griffin et al., 2010, Griffin et al., 2014, Griffin et al., 2016, White et al., 2018).

### Data sources

We calibrated the model to baseline transmission intensity in all malaria-endemic countries before scale-up of interventions, using the year 2000 as an indicator of these levels in line with the current allocation approach taken by the Global Fund (The Global Fund, 2019). Annual EIR was used as a measure of parasite transmission intensity, representing the rate at which people are bitten by infectious mosquitoes. We simulated models to represent a wide range of EIRs for *P. falciparum* and *P. vivax*. These transmission settings were matched to 2000 country-level prevalence data resulting in EIRs of 0.001-80 for *P. falciparum* and 0.001-1.3 for *P. vivax. P. falciparum* estimates came from parasite prevalence in children aged 2-10 years and *P. vivax* prevalence estimates came from light microscopy data across all ages, based on standard reporting for each species (Weiss et al., 2019, Battle et al., 2019). The relationship between parasite prevalence and EIR for specific countries is shown in **Figures S5 and S6**. In each country, the population at risk for *P. falciparum* and *P. vivax* malaria was obtained by summing WorldPop gridded 2000 global population estimates (Tatem, 2017) within Malaria Atlas Project transmission spatial limits using geoboundaries (Runfola et al., 2020) (Supplementary Materials: Country-level data). The analysis was conducted on the national level, since this scale also applies to funding decisions made by international donors (The Global Fund, 2019). As this exercise represents a simplification of reality, population sizes were held constant, and projected population growth is not reflected in the number of cases and the population at risk in different settings. Seasonality was also not incorporated in the model, as EIRs are matched to annual prevalence estimates and the effects of seasonal changes are averaged across the time frame captured. For all analyses, countries were grouped according to their EIR, resulting in a range of transmission settings compatible with the global distribution of malaria. Results were further summarized by grouping EIRs into broader transmission intensity settings according to WHO prevalence cut-offs of 0-1%, 1-10% 10-35% and ≥35% (World Health Organization, 2022a). This corresponded approximately to classifying EIRs of less than 0.1, 0.1 to 1, 1 to 7 and 7 or higher as very low, low, moderate and high transmission intensity, respectively.

### Interventions

In all transmission settings, we simulated the impact of varying coverages of ITNs on clinical incidence. While most countries implement a package of combined interventions, to reduce the computational complexity of the optimization we considered the impact of ITN usage alone in addition to 40% treatment of clinical disease. ITNs are a core intervention recommended for large-scale deployment in areas with ongoing malaria transmission by WHO (Winskill et al., 2019, World Health Organization, 2022a) and funding for vector control represents much of global investments required for malaria control and elimination (Patouillard et al., 2017). Modelled coverages represent population ITN usage between 0 and 80%, with the upper limit reflective of common targets for universal access (Koenker et al., 2018). In each setting, the models were run until clinical incidence stabilized at a new equilibrium with the given ITN usage.

Previous studies have shown that, as population coverage of ITNs increases, the marginal cost of distribution increases as well (Bertozzi-Villa et al., 2021). We incorporated this non-linearity in costs by estimating the annual ITN distribution required to achieve the simulated population usage based on published data from across Africa, assuming that nets would be distributed on a 3-yearly cycle and accounting for ITN retention over time (Supplementary Material). The cost associated with a given simulated ITN usage was calculated by multiplying the number of nets distributed per capita per year by the population size and by the unit cost of distributing an ITN, assumed to be $3.50 (Sherrard-Smith et al., 2022).

### Optimization

The optimal funding allocation for case reduction was determined by finding the allocation of ITNs *b* across transmission settings that minimizes the total number of malaria cases at equilibrium. Case totals were calculated as the sum of the product of clinical incidence *cinc_i_* and the population *p_i_* in each transmission setting *i*. Simultaneous optimization for case reduction and pre-elimination was implemented with an extra weighting term in the objective function, corresponding to a reduction in total remaining cases by a proportion *w* of the total cases averted by the ITN allocation, *C*. This therefore represents a positive contribution for each setting reaching the pre-elimination phase. The weighting on pre-elimination compared to case reduction was 0 in the scenario optimized for case reduction, and varied between 0.5 and 100 times in the other optimization scenarios. Resource allocation must respect a budget constraint, which requires that the sum of the cost of the ITNs distributed cannot exceed the initial budget *B*, with *b_i_* the initial number of ITNs distributed in setting *i* and *c* the cost of a single pyrethroid-treated net. The second constraint requires that the ITN usage 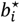 must be between 0 and 80% (Koenker et al., 2018), with ITN usage being a function of ITNs distributed, as shown in the following equation.

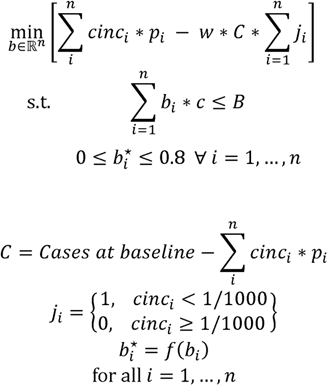

The optimization was undertaken using generalized simulated annealing (Xiang et al., 2013). We included a penalty term in the objective function to incorporate linear constraints. Further details can be found in the Supplementary Material.

The optimal allocation strategy for minimizing cases was also examined over a period of 39 years using the *P. falciparum* model, comparing a single allocation of a constant ITN usage to minimize clinical incidence at 39 years, to reallocation every 3 years (similar to Global Fund allocation periods (The Global Fund, 2016)) leading to varying ITN usage over time. At the beginning of each 3-year period, we determined the optimized allocation of resources to be held fixed until the next round of funding, with the objective of minimizing 3-year global clinical incidence. Once *P. falciparum* elimination is reached in a given setting, ITN distribution is discontinued, and in the next period the same total budget *B* will be distributed among the remaining settings. We calculated the total budget required to minimize case numbers at 39 years and compared the impact of re-allocating each 3 years with a one-time allocation of 25%, 50%, 75% and 100% of the budget. To ensure computational feasibility, 39 years was used as it was the shortest time frame over which the effect of re-distribution of funding from countries having achieved elimination could be observed.

### Analysis

We compared the impact of the two optimal allocation strategies (scenario 1A and 1B) and three additional allocation scenarios on global malaria cases and the global population at risk. Modelled scenarios are shown in **Table 2**. Scenarios 1C-1E represent existing policy strategies that involve prioritizing high-transmission settings, prioritizing low-transmission (near-elimination) settings, or resource allocation proportional to disease burden in the year 2000. Global malaria case burden and the population at risk were compared between baseline levels in 2000 and after reaching an endemic equilibrium under each scenario for a given budget.

**Table 2.**
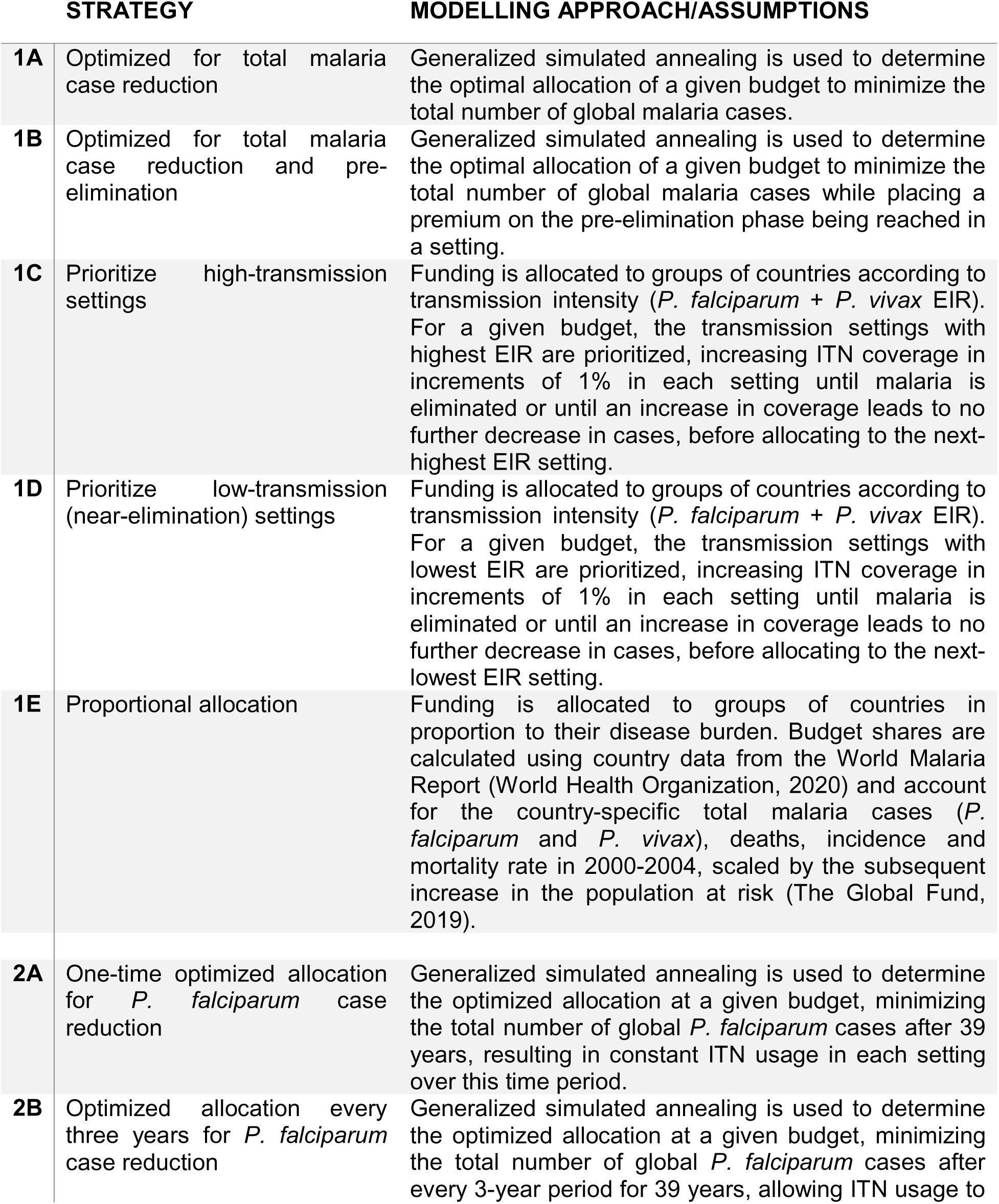
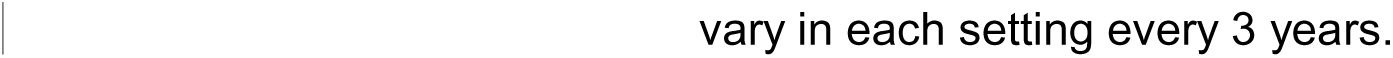
Overview of modelled scenarios for allocation of funding to different transmission settings. Strategies 1A-1E compare resource allocation scenarios using clinical incidence values from each transmission setting at equilibrium after ITN coverage has been introduced. Strategies 2A-2B are compared as part of the allocation over time sub-analysis. EIR: entomological inoculation rate.

| STRATEGY |  | MODELLING APPROACH/ASSUMPTIONS |
| --- | --- | --- |
| 1A | Optimized for total malaria case reduction | Generalized simulated annealing is used to determine the optimal allocation of a given budget to minimize the total number of global malaria cases. |
| 1B | Optimized for total malaria case reduction and pre-elimination | Generalized simulated annealing is used to determine the optimal allocation of a given budget to minimize the total number of global malaria cases while placing a premium on the pre-elimination phase being reached in a setting. |
| 1C | Prioritize high-transmission settings | Funding is allocated to groups of countries according to transmission intensity (*P. falciparum* + *P. vivax* EIR). For a given budget, the transmission settings with highest EIR are prioritized, increasing ITN coverage in increments of 1% in each setting until malaria is eliminated or until an increase in coverage leads to no further decrease in cases, before allocating to the next-highest EIR setting. |
| 1D | Prioritize low-transmission (near-elimination) settings | Funding is allocated to groups of countries according to transmission intensity (*P. falciparum* + *P. vivax* EIR). For a given budget, the transmission settings with lowest EIR are prioritized, increasing ITN coverage in increments of 1% in each setting until malaria is eliminated or until an increase in coverage leads to no further decrease in cases, before allocating to the next-lowest EIR setting. |
| 1E | Proportional allocation | Funding is allocated to groups of countries in proportion to their disease burden. Budget shares are calculated using country data from the World Malaria Report (World Health Organization, 2020) and account for the country-specific total malaria cases (*P. falciparum* and *P. vivax*), deaths, incidence and mortality rate in 2000-2004, scaled by the subsequent increase in the population at risk (The Global Fund, 2019). |
| 2A | One-time optimized allocation for *P. falciparum* case reduction | Generalized simulated annealing is used to determine the optimized allocation at a given budget, minimizing the total number of global *P. falciparum* cases after 39 years, resulting in constant ITN usage in each setting over this time period. |
| 2B | Optimized allocation every three years for *P. falciparum* case reduction | Generalized simulated annealing is used to determine the optimized allocation at a given budget, minimizing the total number of global *P. falciparum* cases after every 3-year period for 39 years, allowing ITN usage to |

Certification of malaria elimination requires proof that the chain of indigenous malaria transmission has been interrupted for at least 3 years and a demonstrated capacity to prevent return transmission (World Health Organization, 2018). In our analysis, transmission settings were defined as having reached malaria elimination once less than one case remained per the setting’s total population. Once a setting reaches elimination, the entire population is removed from the global total population at risk, representing a ‘shrink the map’ strategy. The pre-elimination phase was defined as having reached less than 1 case per 1000 persons at risk in a setting (Mendis et al., 2009).

All strategies were evaluated at different budgets ranging from 0 to the minimum investment required to achieve the lowest possible number of cases in the model (noting that ITNs alone are not predicted to eradicate malaria in our model). No distinctions were made between national government spending and international donor funding, as the purpose of the analysis was to look at resource allocation and not to recommend specific internal and external funding choices.

All analyses were conducted in R v. 4.0.5 (R Foundation for Statistical Computing, Vienna, Austria). The sf (v. 0.9-8, Pebesma 2018), raster (v. 3.4-10, Hijmans & van Etten 2012), and terra (v.1.3-4, Hijmans 2021) packages were used for spatial data manipulation. The akima package (v.0.6-2.2, Akima and Gebhardt 2021) was used for surface development, and the GenSA package (v.1.1.7, Gubian et al.) for model optimization.

## AUTHOR CONTRIBUTIONS

N. S, H. M. T., M. P., G. D. C., K. H. and A. C. G. conceived the study. N. S., H. M. T., and M. P. processed the data, conducted the analysis, and created the visualizations. N. S., H. M. T., M. P. and G. D. C. developed the computational infrastructure and methodology. P. W., M. T. W., K. H. and A. C. G. contributed domain knowledge. K. H. and A. C. G. acquired funding for the study and provided guidance on analysis and interpretation. N. S. and H. M. T. wrote the first draft of the manuscript. All authors contributed to interpreting the results, reviewing and revising the manuscript, and approved the final version for submission.

## Data Availability

Datasets of parasite prevalence and spatial limits are publicly available from the Malaria Atlas Project at https://malariaatlas.org/. The previously published malaria transmission models code is available to download at https://github.com/mrc-ide/deterministic-malaria-model. The code to conduct the analysis and produce the figures and tables in the manuscript are available to download at https://github.com/mrc-ide/malaria_optimal_allocation.

