## Supplementary Materials for "Resource allocation strategies for insecticide-treated bed nets to achieve malaria eradication"

#### Supplementary Material

Nora Schmit[1]*, Hillary M Topazian[1]*, Matteo Pianella[1]*, Giovanni D Charles[1], Peter Winskill[1], Michael T White[2], Katharina Hauck[1], Azra C Ghani[1]

[1] MRC Centre for Global Infectious Disease Analysis, Imperial College London, London, United Kingdom

[2] Infectious Disease Epidemiology and Analytics G5 Unit, Department of Global Health, Institut Pasteur, Université de Paris, Paris, France

* These authors contributed equally to this work

### SUPPLEMENTARY METHODS

#### Mathematical models

##### 1. Overview

In this paper we use an existing deterministic, compartmental, mathematical model of *P. falciparum* malaria transmission between humans and mosquitoes, which was originally calibrated to age-stratified data from settings across sub-Saharan Africa (1). We also developed a deterministic version of an existing individual-based model of *P. vivax* transmission, originally calibrated to data from Papua New Guinea but also shown to reproduce global epidemiological patterns (2). Both models are structured by age and heterogeneity in exposure to mosquito bites, and allow for the presence of maternal immunity at birth and naturally acquired immunity across the life course. The mosquito and vector control components are modelled identically in both models, except for the force of infection acting on mosquitoes. A diagram of the model structures with human and adult mosquito components is shown in **Figure S1**.

Population and transmission dynamics were modelled separately for both species, assuming they are independent of each other, because the epidemiological significance of biological interactions between the parasites within hosts remains unclear (3).

Note that while the term ‘individuals’ may be used in descriptions, the models are compartmental and do not track individuals; compartments represent the average number of people in a given state.

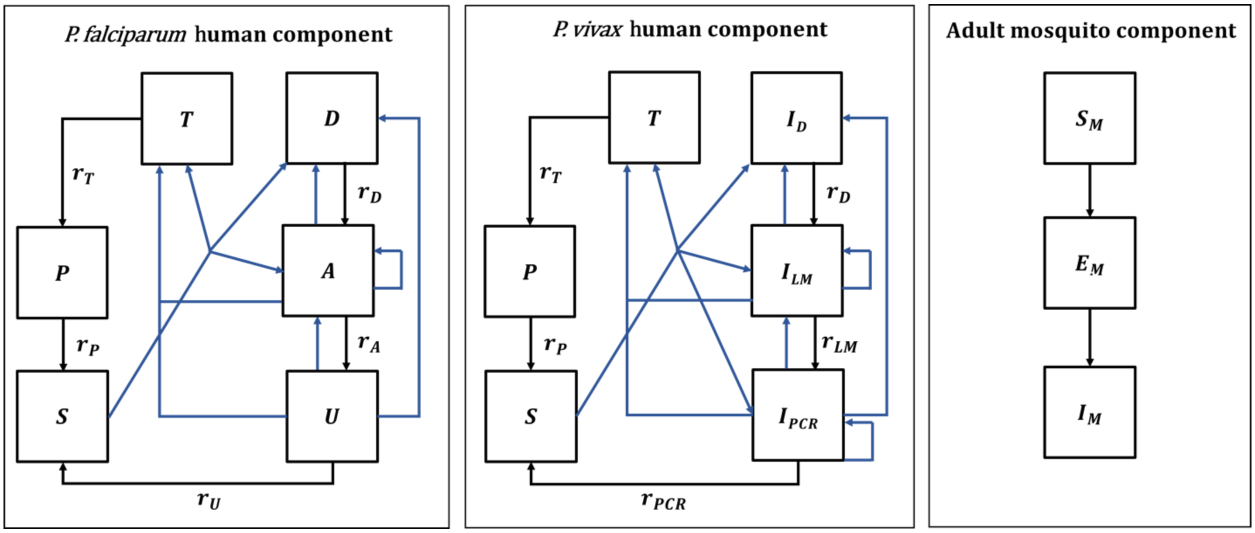


**Figure S1**. Malaria transmission model; diagram adapted from Griffin et al. 2016 and White et al. 2018 (1, 2). Humans move through six states in the models for both species: $S$ (susceptible), $D$ (untreated symptomatic infection), $T$ (successfully treated symptomatic infection), $A$ (asymptomatic infection), $U$ (asymptomatic sub-patent infection) and $P$ (prophylaxis) in the *P. falciparum* model, and $S$ (susceptible), $I_D$ (untreated symptomatic infection), $T$ (successfully treated symptomatic infection), $I_{LM}$ (asymptomatic light microscopy detectable blood-stage infection), $I_{PCR}$ (asymptomatic sub-microscopic PCR detectable blood-stage infection) and $P$ (prophylaxis) in the *P. vivax* model. New infections (including superinfections) are highlighted in blue but parameters are not shown. Rates $r_D$, $r_A$, $r_U$, $r_T$, $r_P$, $r_{LM}$, and $r_{PCR}$ determine the mean duration of each state. Hypnozoites states in the *P. vivax* model are not shown on the diagram. Adult female mosquitoes move through three model compartments: $S_m$ (susceptible), $E_m$ (exposed), and $I_m$ (infected).

#### 2. Human demography

In both the *P. falciparum* and *P. vivax* model, the aging process in the human population follows an exponential distribution. Humans can reach a maximum age of 100 years and experience a constant death rate of 1/21 per year based on the assumed median age of the population. The birth rate was assumed to equal the mortality rate so that the population remains stable over time. Demographic changes over time are therefore not accounted for.

In all following sections, human demographic processes are omitted from the equations of the transmission models and the immunity models for simplicity. All compartments experience the same constant mortality rate, while all births occur in the susceptible compartment.

#### 3. Heterogeneity in mosquito biting rates

In both models, exposure to mosquito bites is assumed to depend on age, due to varying body surface area and behavioral patterns.

The relative biting rate at age $a$ is calculated as:

$$\psi(a) = (1 - \rho \exp(-a/a_0)) \tag{3.1}$$

Where $\rho$ and $a_0$ are estimated parameters determining the relationship between age and biting rate.

Additionally, the human population in the model is stratified according to lifetime relative biting rate $\zeta$which represents the heterogeneity in exposure to mosquito bites that occurs at various spatial scales, e.g. due to attractiveness of humans to mosquitoes, housing standards and proximity to mosquito breeding sites, and is described by a log-normal distribution with a mean of 1, as follows:

$$\log(\zeta) \sim N(-\sigma^2/2\,, \sigma^2) \tag{3.2}$$

#### 4. *P. falciparum* human model component

In the *P. falciparum* model, humans move through four states of transmission and are present in only one of the six states at each timestep: susceptible ($S$), untreated symptomatic infection ($D$), successfully treated symptomatic infection ($T$), asymptomatic infection ($A$), asymptomatic sub-patent infection ($U$) and prophylaxis ($P$). Individuals in the model are born susceptible to infection but are temporarily protected by maternal immunity during the first six months of life. Humans are exposed to infectious bites from mosquitoes and are infected at a rate $\Lambda$, representing the force of infection from mosquitoes to humans. The force of infection depends on an individual's pre-erythrocytic immunity, the age-dependent biting rate and the mosquito population size and level of infectivity. Following a latent period, $d_E$, and depending on clinical immunity levels, a proportion $\phi$ of infected individuals develop clinical disease, while the remaining move into the asymptomatic infection state. A proportion $f_T$ of those with clinical disease are successfully treated. Treated individuals recover from infection at rate $r_T$ and move to the prophylaxis state, which represents a period of drug-dependent partial protection from reinfection. Recovery from untreated symptomatic infection to the asymptomatic infection state occurs at a rate $r_D$, while those with asymptomatic infection develop sub-patent infection at a rate $r_A$. The sub-patent infection and prophylaxis states then clear infection and return to the susceptible state at rates $r_U$ and $r_P$, respectively. In the susceptible compartment, re-infection can occur, while asymptomatic and sub-patent infection are also susceptible to superinfection, potentially giving rise to further clinical cases. *P. falciparum* parameters are listed in **Table S1**.

The human component of the model is described by the following set of partial differential equations with regards to time $t$ and age $a$:

$$\frac{\partial S}{\partial t} + \frac{\partial S}{\partial a} = -\Lambda(t - d_E)S + r_U U + r_P P$$

$$\frac{\partial D}{\partial t} + \frac{\partial D}{\partial a} = \phi(1 - f_T)\Lambda(t - d_E)(S + A + U) - r_D D$$

$$\frac{\partial T}{\partial t} + \frac{\partial T}{\partial a} = \phi f_T \Lambda(t - d_E)(S + A + U) - r_T T \quad (4.1)$$

$$\frac{\partial A}{\partial t} + \frac{\partial A}{\partial a} = (1 - \phi)\Lambda(t - d_E)(S + U) + r_D D - \phi\Lambda(t - d_E)A - r_A A$$

$$\frac{\partial U}{\partial t} + \frac{\partial U}{\partial a} = r_A A - \Lambda(t - d_E)U - r_U U$$

$$\frac{\partial P}{\partial t} + \frac{\partial P}{\partial a} = r_T T - r_P P$$

Note that age- and time-dependence in state variables and parameters, as well as mortality and birth rates, are omitted in equations for clarity.

Accounting for the heterogeneity and age-dependence in mosquito biting rates described above, the force of infection $\Lambda(a,t)$ and the EIR (entomological inoculation rate) $\varepsilon(a,t)$ for age $a$ at time $t$ are given by:

$$\Lambda(a,t) = \varepsilon(a,t)b \quad (4.2)$$

$$\varepsilon(a,t) = \varepsilon_0(t)\zeta\psi(a) \quad (4.3)$$

Where $\varepsilon_0$ is the mean entomological inoculation rate (EIR) experienced by adults at time $t$, and $b$ is the probability that a human will be infected when bitten by an infectious mosquito.

The mean EIR experienced by adults is represented by:

$$\varepsilon_0(t) = \frac{\alpha I_M}{\omega} \quad (4.4)$$

Where $\alpha$ is the mosquito biting rate on humans, $I_M$ is the compartment for adult

infectious mosquitoes (see vector model component), and $\omega$ is a normalization constant for the biting rate over various age groups with a population age distribution of $\eta(a)$, as follows.

$$\omega = \int_0^\infty \eta(a)\psi(a)da \quad (4.5)$$

The probability of infection $b$, probability of clinical symptomatic disease $\phi$, and recovery rate from asymptomatic infection $r_A$, all depend on immunity levels. The acquisition and decay of naturally-acquired immunity is tracked dynamically in the model and is driven by both age and exposure. Naturally-acquired immunity affects three different outcomes in the model, leading to: 1) a reduced probability of developing a blood-stage infection following an infectious bite due to pre-erythrocytic immunity, $I_B$, 2) a reduced probability of progression to clinical disease following infection, dependent on exposure-driven and maternally acquired clinical immunity, $I_{CA}$ and $I_{CM}$, and 3) a reduced probability of a blood-stage infection being detected by microscopy, dependent on acquired immunity to the detectability of infection, $I_D$.

The following partial differential equations represent exposure-driven immunity levels at time $t$ and age $a$.

Pre-erythrocytic immunity: $$\frac{\partial I_B}{\partial t} + \frac{\partial I_B}{\partial a} = \frac{\varepsilon}{\varepsilon u_B + 1} - \frac{I_B}{d_B} \quad (4.6)$$

Clinical immunity: $$\frac{\partial I_{CA}}{\partial t} + \frac{\partial I_{CA}}{\partial a} = \frac{\Lambda}{\Lambda u_C + 1} - \frac{I_{CA}}{d_{CA}} \quad (4.7)$$

Detection immunity: $$\frac{\partial I_D}{\partial t} + \frac{\partial I_D}{\partial a} = \frac{\Lambda}{\Lambda u_D + 1} - \frac{I_D}{d_{ID}} \quad (4.8)$$

Where $u$ parameters represent a refractory period during which the different types of immunity cannot be further boosted after receiving a boost, and where $d$ parameters stand for the mean duration of the different types of immunity.

Maternal immunity is acquired and lost as follows:

$$\frac{\partial I_{CM}}{\partial t} + \frac{\partial I_{CM}}{\partial a} = -\frac{I_{CM}}{d_{CM}} \quad (4.9)$$

$$I_{CM}(t, 0) = P_{CM} I_{CA}(t, 20)$$

Where $d_{CM}$ is the average duration of maternal immunity, $P_{CM}$ is the proportion of the mother's clinical immunity acquired by the newborn, and $I_{CA}(t, 20)$ denotes the clinical immunity level of a 20-year-old woman.

Immunity levels are converted into time- and age-dependent probabilities using Hill functions.

The probability that a human will be infected when bitten by an infectious mosquito, $b$, can be represented as:

$$b = b_0\left(b_1 + \frac{1-b_1}{1+(I_B/I_{B0})^{\kappa_B}}\right) \tag{4.10}$$

Where $b_0$ is the maximum probability of infection (with no immunity), $b_1$ is the maximum relative reduction in the probability of infection due to immunity, and $I_{B0}$ and $\kappa_B$ are scale and shape parameters estimated during model fitting.

The probability of a new blood-stage infection becoming symptomatic, $\phi$, is represented by:

$$\phi = \phi_0\left(\phi_1 + \frac{1-\phi_1}{1+((I_{CA}+I_{CM})/I_{C0})^{\kappa_C}}\right) \tag{4.11}$$

Where $\phi_0$ is the maximum probability of becoming symptomatic (with no immunity), $\phi_1$ is the maximum relative reduction in the probability of becoming symptomatic due to immunity, and $I_{C0}$ and $\kappa_C$ are scale and shape parameters, respectively.

Immunity can also lead to blood-stage infections becoming sub-patent with low parasitemias. The probability that an asymptomatic infection is detectable by microscopy, $q$, is represented by:

$$q = d_1 + \frac{1-d_1}{1+f_D(I_D/I_{D0})^{\kappa_D}} \tag{4.12}$$

Where $d_1$ is the minimum probability of detectability (with full immunity), and $I_{D0}$ and $\kappa_D$ are scale and shape parameters, respectively. $f_D$ is an age-dependent function modifying the detectability of infection:

$$f_D = 1 - \frac{1-f_{D0}}{1+\left(\frac{a}{a_D}\right)^{\gamma_D}} \tag{4.13}$$

With $\gamma_D$ and $a_D$ representing shape and scale parameters, and $f_{D0}$ representing the time-scale at which immunity changes with age.

**Table S1**. *P. falciparum* human model parameter values. Full details can be found in the original publication (1), including references for parameters and intervals for the prior and posterior distributions (median values of the posterior distribution are used in model simulations).

| Parameter | Symbol | Estimate |
|---|---|---|
| **Human infection duration (days)** | | |
| Latent period | $d_E$ | 12 |
| Patent infection | $1/r_A$ | 195 |
| Clinical disease (untreated) | $1/r_D$ | 5 |
| Treatment of clinical disease | $1/r_T$ | 5 |
| Sub-patent infection | $1/r_U$ | 110.299 |
| Prophylaxis | $1/r_P$ | 15 |
| **Age and heterogeneity** | | |
| Age-dependent biting parameter | $\rho$ | 0.85 |
| Age-dependent biting parameter | $a_0$ | 8 years |
| Variance of the log heterogeneity in biting rates | $\sigma^2$ | 1.67 |
| **Pre-erythrocytic immunity reducing probability of infection** | | |
| Duration of refractory period in which immunity is not boosted | $u_B$ | 7.19919 days |
| Duration of pre-erythrocytic immunity | $d_B$ | 10 years |
| Maximum probability of infection due to no immunity | $b_0$ | 0.590076 |
| Maximum relative reduction in probability of infection due to immunity | $b_1$ | 0.5 |
| Scale parameter | $I_{B0}$ | 43.8787 |
| Shape parameter | $K_B$ | 2.15506 |
| **Immunity reducing probability of clinical disease** | | |
| Duration of refractory period in which immunity is not boosted | $u_C$ | 6.06349 days |
| Duration of clinical immunity | $d_{CA}$ | 30 years |
| New-born immunity relative to mother's clinical immunity | $P_{CM}$ | 0.774368 |
| Duration of maternal immunity | $d_{CM}$ | 67.6952 days |
| Maximum probability of clinical disease due to no immunity | $\Phi_0$ | 0.791666 |
| Maximum relative reduction in probability of clinical disease due to immunity | $\Phi_1$ | 0.000737 |
| Scale parameter | $I_{C0}$ | 18.02366 |
| Shape parameter | $K_C$ | 2.36949 |
| **Immunity reducing probability of detection** | | |
| Duration of refractory period in which immunity is not boosted | $u_D$ | 9.44512 days |
| Duration of detection immunity | $d_{ID}$ | 10 years |
| Minimum probability of detection due to maximum immunity | $d_1$ | 0.160527 |
| Scale parameter | $I_{D0}$ | 1.577533 |
| Shape parameter | $K_D$ | 0.476614 |
| Scale parameter relating age to immunity | $a_D$ | 21.9 years |
| Time-scale at which immunity changes with age | $f_{D0}$ | 0.007055 |
| Shape parameter relating age to immunity | $\gamma_D$ | 4.8183 |

#### 5. *P. vivax* human model component

In the *P. vivax* model, acquisition, and recovery from blood-stage infection in the absence of treatment is also represented by four compartments: susceptible ($S$), untreated symptomatic infection ($I_D$), successfully treated symptomatic infection ($T$), asymptomatic patent blood-stage infection detectable by light microscopy ($I_{LM}$), asymptomatic sub-microscopic infection not detectable by light microscopy, but detectable by PCR ($I_{PCR}$), and prophylaxis ($P$). Additionally, the model represents the liver stage of *P. vivax* infection by tracking average hypnozoite batches in the population. Hypnozoites can form after an infectious bite and remain dormant in the liver for up to several years, which can give rise to relapse blood-stage infections. *P. vivax* parameters are listed in **Table S2**.

New blood-stage infections can therefore originate from either mosquito bites or relapses and are represented by the force of infection $\lambda_H^0$. The force of infection depends on the age-dependent biting rate, the mosquito population size and its level of infectivity, the probability of infection resulting from an infectious bite, the latent period between sporozoite inoculation and development of blood-stage merozoites, $d_E$, as well as relapse infections from the liver stage. Upon infection, a proportion $\Phi_{LM}$ of humans develop infection detectable by light microscopy (LM), while the remainder have low-density parasitemia and move into the $I_{PCR}$ compartment. A proportion $\Phi_D$ of those with LM-detectable infection develop a clinical episode, of which a proportion $X_T$ are successfully treated with a blood-stage antimalarial. Treated individuals recover from infection at rate $r_T$ and move to the prophylaxis state, which provides temporary protection from reinfection before becoming susceptible again at a rate $r_P$. Recovery from clinical disease to asymptomatic LM-detectable infection, from asymptomatic LM-detectable infection to asymptomatic PCR-detectable infection and from asymptomatic PCR-detectable infection to susceptibility occur at rates $r_D$, $r_{LM}$ and $r_{PCR}$, respectively. Newborns are susceptible to infection, have no hypnozoites and are temporarily protected by maternal immunity. Reinfection is possible after recovery, and those with asymptomatic blood stage infections ($I_{LM}$ and $I_{PCR}$) are susceptible to superinfection, potentially giving rise to further clinical cases.

The dynamics of hypnozoite infection in the model describe the accumulation and clearance of $k$ batches of hypnozoites in the liver, whereby each new (super-)infection from an infectious mosquito bite creates a new batch. This process occurs for each model compartment and is described in detail in the original publication (2). Hypnozoites from any batch can re-activate and cause a relapse at a rate $kf$, and batches are cleared at a constant rate $k\gamma_L$, which reduces the number of batches from $k$ to $k-1$. For computational efficiency, the possible number of batches in the population must be limited to a maximum value $K$, so that superinfections among the population with $k = K$ do not lead to an increase in hypnozoite batch numbers. We assumed a maximum batch number of 2, which increased computational efficiency and aligned with modelled distributions of hypnozoite batch numbers in the population for the simulated low transmission intensities.

The human component of the model is then described by the following set of partial differential equations with regards to time $t$ and age $a$:

$$
\begin{aligned}
\frac{\partial S^k}{\partial t}+\frac{\partial S^k}{\partial a} &= -\lambda_H^0(t-d_E)S^k - fkS^k + r_{PCR}^k I_{PCR}^k - \gamma_L k S^k + \gamma_L(k+1)S^{k+1} \\
\frac{\partial I_{PCR}^k}{\partial t}+\frac{\partial I_{PCR}^k}{\partial a} &= -\lambda_H^0(t-d_E)I_{PCR}^k - fkI_{PCR}^k - r_{PCR}^k I_{PCR}^k + r_{LM}I_{LM}^k \\
&\quad + \lambda_H^0(t-d_E)\left(1-\Phi_{LM}^{k-1}\right)(S^{k-1}+I_{PCR}^{k-1}) + fk\left(1-\Phi_{LM}^k\right)(S^k+I_{PCR}^k) \\
&\quad - \gamma_L k I_{PCR}^k + \gamma_L(k+1)I_{PCR}^{k+1} \\
\frac{\partial I_{LM}^k}{\partial t}+\frac{\partial I_{LM}^k}{\partial a} &= -\lambda_H^0(t-d_E)I_{LM}^k - fkI_{LM}^k - r_{LM}I_{LM}^k + r_D I_D^k \\
&\quad + \lambda_H^0(t-d_E)\left(1-\Phi_D^{k-1}\right)(\Phi_{LM}^{k-1}S^{k-1} + \Phi_{LM}^{k-1}I_{PCR}^{k-1} + I_{LM}^{k-1}) \\
&\quad + fk\left(1-\Phi_D^k\right)(\Phi_{LM}^k S^k + \Phi_{LM}^k I_{PCR}^k + I_{LM}^k) - \gamma_L k I_{LM}^k + \gamma_L(k+1)I_{LM}^{k+1} \\
\frac{\partial I_D^k}{\partial t}+\frac{\partial I_D^k}{\partial a} &= -\lambda_H^0(t-d_E)I_D^k + \lambda_H^0(t-d_E)I_D^{k-1} - r_D I_D^k + \lambda_H^0(t-d_E)\Phi_D^{k-1}(1 \\
&\quad - X_t)(\Phi_{LM}^{k-1}S^{k-1} + \Phi_{LM}^{k-1}I_{PCR}^{k-1} + I_{LM}^{k-1}) + fk\Phi_D^k(1-X_t)(\Phi_{LM}^k S^k \\
&\quad + \Phi_{LM}^k I_{PCR}^k + I_{LM}^k) - \gamma_L k I_D^k + \gamma_L(k+1)I_D^{k+1} \\
\frac{\partial T^k}{\partial t}+\frac{\partial T^k}{\partial a} &= -\lambda_H^0(t-d_E)T^k + \lambda_H^0(t-d_E)T^{k-1} - r_T T^k \\
&\quad + \lambda_H^0(t-d_E)\Phi_D^{k-1}X_t(\Phi_{LM}^{k-1}S^{k-1} + \Phi_{LM}^{k-1}I_{PCR}^{k-1} + I_{LM}^{k-1}) \\
&\quad + fk\Phi_D^k X_t(\Phi_{LM}^k S^k + \Phi_{LM}^k I_{PCR}^k + I_{LM}^k) - \gamma_L k T^k + \gamma_L(k+1)T^{k+1} \\
\frac{\partial P^k}{\partial t}+\frac{\partial P^k}{\partial a} &= -\lambda_H^0(t-d_E)P^k + \lambda_H^0(t-d_E)P^{k-1} + r_T T^k - r_P P^k - \gamma_L k P^k \\
&\quad + \gamma_L(k+1)P^{k+1}
\end{aligned} \tag{5.1}
$$

Where $fk$ and $\gamma_L k$ are the relapse and clearance rates of hypnozoite batch $k$, respectively. Age- and time-dependence in state variables and parameters, as well as mortality and birth rates, are omitted in the equations for clarity.

The equations reflect the accumulation of hypnozoite batches from $k$ to $k+1$ due to infections arising from new infectious bites ($\lambda_H^0$), but not due to relapse infections ($fk$). The total force of blood-stage infection is therefore:

$$
\lambda_H^k(t-d_E) = \lambda_H^0(t-d_E) + kf \tag{5.2}
$$

Similar to the *P. falciparum* model, the force of infection from mosquito bites accounts for heterogeneity and age-dependence in mosquito biting rates as follows:

$$
\lambda_H^0(a,t) = \varepsilon(a,t)b \tag{5.3}
$$

$$
\varepsilon(a,t) = \varepsilon_0(t)\zeta\psi(a) \tag{5.4}
$$

$$
\varepsilon_0(t) = \frac{\alpha I_M}{\omega} \tag{5.5}
$$

$$
\omega = \int_0^\infty \eta(a)\psi(a)\,da \tag{5.6}
$$

Where $\varepsilon_0$ is the mean entomological inoculation rate (EIR) experienced by adults at time $t$, and $b$ is the probability that a human will be infected when bitten by an infectious mosquito. In the *P. vivax* model, $b$ is a constant and does not depend on immunity levels. In the calculation of the mean EIR experienced by adults, $\alpha$ is the mosquito biting rate on humans, $I_M$ is the compartment for adult infectious mosquitoes (see vector model component), and $\omega$ is a normalization constant for the biting rate over various age groups with a population age distribution of $\eta(a)$.

Transmission dynamics in the model are influenced by anti-parasite ($A_P$) and clinical immunity ($A_C$) against *P. vivax*. Anti-parasite immunity is assumed to reduce the probability of blood-stage infections achieving high enough density to be detectable by light microscopy ($\Phi_{LM}$) and to increase the rate at which sub-microscopic infections are cleared ($r_{PCR}$). Clinical immunity reduces the probability that LM-detectable infections progress to clinical disease ($\Phi_D$). Like for *P. falciparum*, the dynamics of the acquisition and decay of naturally-acquired immunity in the model depend on age and exposure. For *P. vivax*, immunity levels are boosted by both primary infections and relapses and are described by the following set of partial differential equations with regards to time $t$ and age $a$:

Anti-parasite immunity:

$$\frac{\partial A_P^0}{\partial t} + \frac{\partial A_P^0}{\partial a} = -\lambda_H^0(t-d_E)A_P^0 - r_{par}A_P^0 + \gamma_L A_P^1$$

$$\frac{\partial A_P^k}{\partial t} + \frac{\partial A_P^k}{\partial a} = \frac{\lambda_H^k(t-d_E)}{\lambda_H^k(t-d_E)u_{par}+1} - \lambda_H^0(t-d_E)A_P^k + \lambda_H^0(t-d_E)A_P^{k-1} - r_{par}A_P^k - \gamma_L k A_P^k + \gamma_L(k+1)A_P^{k+1} \tag{5.7}$$

$$\frac{\partial A_P^K}{\partial t} + \frac{\partial A_P^K}{\partial a} = \frac{\lambda_H^K(t-d_E)}{\lambda_H^K(t-d_E)u_{par}+1} + \lambda_H^0(t-d_E)A_P^{K-1} - r_{par}A_P^K - \gamma_L K A_P^K$$

Clinical immunity:

$$\frac{\partial A_C^0}{\partial t} + \frac{\partial A_C^0}{\partial a} = -\lambda_H^0(t-d_E)A_C^0 - r_C A_C^0 + \gamma_L A_C^1$$

$$\frac{\partial A_C^k}{\partial t} + \frac{\partial A_C^k}{\partial a} = \frac{\lambda_H^k(t-d_E)}{\lambda_H^k(t-d_E)u_C+1} - \lambda_H^0(t-d_E)A_C^k + \lambda_H^0(t-d_E)A_C^{k-1} - r_C A_C^k - \gamma_L k A_C^k + \gamma_L(k+1)A_C^{k+1} \tag{5.8}$$

$$\frac{\partial A_C^K}{\partial t} + \frac{\partial A_C^K}{\partial a} = \frac{\lambda_H^K(t-d_E)}{\lambda_H^K(t-d_E)u_C+1} + \lambda_H^0(t-d_E)A_C^{K-1} - r_C A_C^K - \gamma_L K A_C^K$$

Where $u$ parameters represent a refractory period during which the different types of immunity cannot be further boosted after receiving a boost, and where $r$ parameters stand for the rates of decay of the different types of immunity. $k$ refers to the hypnozoite batch (with $K$ being the maximum number of hypnozoite batches).

The levels of maternally acquired anti-parasite and clinical immunity are calculated as:

$$A_{P,mat}(t,a) = P_{mat}A_P^*(t-a,20)e^{-\frac{a}{d_{mat}}} \quad (5.9)$$

$$A_{C,mat}(t,a) = P_{mat}A_C^*(t-a,20)e^{-\frac{a}{d_{mat}}} \quad (5.10)$$

Where $d_{mat}$ is the average duration of maternal immunity, $P_{mat}$ is the proportion of the mother's immunity acquired by the newborn, and $A_P^*(t-a,20)$ and $A_C^*(t-a,20)$ denote the anti-parasite and clinical immunity level of a 20-year-old woman averaged over their hypnozoite batches, respectively.

Immunity levels are then converted into time-dependent probabilities using Hill functions.

The probability that a blood-stage infection becomes detectable by LM, $\Phi_{LM}$, can be represented as:

$$\Phi_{LM} = \Phi_{LM,min} + (\Phi_{LM,max} - \Phi_{LM,min})\frac{1}{1+\left(\frac{A_P^k + A_{P,mat}}{A_{LM,50\%}}\right)^{K_{LM}}} \quad (5.11)$$

Where $\Phi_{LM,min}$ is the minimum probability of LM-detectable infection (with full immunity), $\Phi_{LM,max}$ is the maximum probability of LM-detectable infection (with no immunity), and $A_{LM,50\%}$ and $K_{LM}$ are scale and shape parameters estimated during model fitting.

The probability of a LM-detectable blood-stage infection becoming symptomatic, $\Phi_D$, is represented by:

$$\Phi_D = \Phi_{D,min} + (\Phi_{D,max} - \Phi_{D,min})\frac{1}{1+\left(\frac{A_C^k + A_{C,mat}}{A_{D,50\%}}\right)^{K_D}} \quad (5.12)$$

Where $\Phi_{D,min}$ is the minimum probability of developing a clinical episode (with full immunity), $\Phi_{D,max}$ is the maximum probability of a clinical episode (with no immunity), and $A_{D,50\%}$ and $K_D$ are scale and shape parameters.

The recovery rate from $I_{PCR}$ is calculated as $\frac{1}{d_{PCR}^k}$. The average duration of a low-density blood-stage infection, $d_{PCR}^k$, is represented by:

$$d_{PCR}^k = d_{PCR,min} + (d_{PCR,max} - d_{PCR,min})\frac{1}{1+\left(\frac{A_P^k + A_{P,mat}}{A_{PCR,50\%}}\right)^{K_{PCR}}} \quad (5.13)$$

Where $d_{PCR,min}$ is the minimum duration (with full immunity), $d_{PCR,max}$ is the maximum duration (with no immunity), and $A_{PCR,50\%}$ and $K_{PCR}$ are scale and shape parameters.

**Table S2**. *P. vivax* human model parameter values. Full details can be found in the original publication (2) including references for parameters and intervals for the prior and posterior distributions.

| Parameter | Symbol | Estimate |
|---|---|---|
| **Human infection duration (days)** | | |
| Latent period | $d_E$ | 10 |
| Light microscopy-detectable asymptomatic infection | $1/r_{LM}$ | 10 |
| Clinical disease (untreated) | $1/r_D$ | 5 |
| Treatment of clinical disease | $1/r_T$ | 1 |
| Prophylaxis | $1/r_P$ | 28 |
| **Age, heterogeneity and probability of infection** | | |
| Age-dependent biting parameter | $\rho$ | 0.85 |
| Age-dependent biting parameter | $a_0$ | 8 years |
| Variance of the log heterogeneity in biting rates | $\sigma^2$ | 1.29 |
| Probability of blood-stage infection upon infectious mosquito bite | $b$ | 0.5 |
| **Hypnozoite parameters** | | |
| Relapse rate | $f$ | 0.024 per day |
| Clearance rate | $\gamma_L$ | 0.0026 per day |
| **Maternal immunity** | | |
| New-born immunity relative to mother's clinical immunity | $P_{mat}$ | 0.421 |
| Duration of maternal immunity | $d_{mat}$ | 35.148 days |
| **Anti-parasite immunity reducing probability of light microscopy-detectable infection and duration of PCR-detectable infection** | | |
| Duration of refractory period in which immunity is not boosted | $u_{par}$ | 19.77 days |
| Duration of anti-parasite immunity | $1/r_{par}$ | 10 years |
| Maximum probability of detectability by light microscopy due to no immunity | $\Phi_{LM,max}$ | 0.8918 |
| Minimum probability of detectability by light microscopy due to full immunity | $\Phi_{LM,min}$ | 0.0043 |
| Scale parameter for detectability by light microscopy | $A_{LM,50\%}$ | 27.52 |
| Shape parameter for detectability by light microscopy | $K_{LM}$ | 2.403 |
| Maximum duration of PCR-detectable infection due to no immunity | $d_{PCR,max}$ | 70 days |
| Minimum duration of PCR-detectable infection due to full immunity | $d_{PCR,min}$ | 10 days |
| Scale parameter for duration of PCR-detectable infection | $A_{PCR,50\%}$ | 9.9 |
| Shape parameter for duration of PCR-detectable infection | $K_{PCR}$ | 4.602 |
| **Clinical immunity reducing probability of clinical disease** | | |
| Duration of refractory period in which immunity is not boosted | $u_C$ | 7.85 days |
| Duration of detection immunity | $1/r_C$ | 30 years |
| Maximum probability of clinical disease due to no immunity | $\Phi_{D,min}$ | 0.8605 |
| Minimum probability of clinical disease due to full immunity | $\Phi_{D,max}$ | 0.018 |
| Scale parameter for clinical disease | $A_{D,50\%}$ | 11.538 |
| Shape parameter for clinical disease | $K_D$ | 2.250 |

#### 6. Mosquito component of the *P. falciparum* and *P. vivax* model

The mosquito components of the *P. falciparum* and *P. vivax* models capture adult mosquito transmission dynamics, as well as larval population dynamics, and nearly identical. Modelled vector bionomics correspond to *Anopheles gambiae s.s.* and *Anopheles punctulatus* for *P. falciparum* and *P. vivax* transmission, respectively.

*Mosquito transmission model*

Adult mosquitoes move between three states, $S_M$ (susceptible), $E_M$ (exposed), and $I_M$ (infectious), as follows:

$$\frac{dS_M}{dt} = -\Lambda_M S_M + \beta(t) - \mu S_M$$
$$\frac{dE_M}{dt} = \Lambda_M S_M - \Lambda_M(t-\tau_M)S_M(t-\tau_M)P_M - \mu E_M \qquad (6.1)$$
$$\frac{dI_M}{dt} = \Lambda_M(t-\tau_M)S_M(t-\tau_M)P_M - \mu I_M$$

$\Lambda_M$ is the force of infection from humans to mosquitos, $\beta(t)$ represents the time-varying adult mosquito emergence rate, $\mu$ is the adult mosquito death rate, and $\tau_M$ represents the extrinsic incubation period. $P_M$ represents the probability that a mosquito survives between being infected and sporozoites appearing in the salivary glands and is calculated as $\exp(-\mu\tau_M)$.

The force of infection experienced by the vector is the sum of the contribution to mosquito infections from all human infectious states. As described for the human model components for both species, it also depends on the mosquito biting rate on humans (which depends on net usage), $\alpha$, and a normalisation constant for the biting rate over various age groups, $\omega$.

*Force of infection experienced by mosquitoes in the P. falciparum model*

In the *P. falciparum* model, the force of infection acting on mosquitoes is represented by:

$$\Lambda_M(t) = \frac{\alpha}{\omega}\iint_{\zeta\alpha} \zeta\psi(a)\big(c_D D(\zeta,a,t-\tau_1) + c_T T(\zeta,a,t-\tau_1) + c_A A(\zeta,a,t-\tau_1) + c_U U(\zeta,a,t-\tau_1)\big)da\, d\zeta \qquad (6.2)$$

Where $c_D$, $c_T$, $c_A$, and $c_U$ represent the human-to-mosquito infectiousness for untreated symptomatic infection, treated symptomatic infection, asymptomatic infection and asymptomatic sub-patent infection, respectively. $\tau_1$ is the time-lag between parasitemia with asexual parasite stages and gametocytemia to account for the time to *P. falciparum* gametocyte development.

The infectiousness of humans with asymptomatic infection, $c_A$, is reduced by a lower probability of detection of infection by microscopy due to the assumption that lower parasite densities are less detectable. While infectiousness parameters $c_D$ and $c_U$ are constant, infectivity for asymptomatic infection is calculated as follows:

$$c_A = c_U + (c_D - c_U)q^{\gamma_1} \qquad (6.3)$$

Where $q$ is the immunity-dependent probability that an asymptomatic infection is detectable by microscopy (equation ( 4.12) and the parameter $\gamma_1$ was estimated during the original model fitting in previous publications (1, 4, 5).

*Force of infection experienced by mosquitoes in the P. vivax model*

In the *P. vivax* model, the force of infection acting on mosquitoes is represented by:

$$\Lambda_M(t) = \frac{\alpha}{\omega} \iint_{\zeta\alpha} \zeta\psi(a)\big(c_D I_D(\zeta,a,t) + c_T T(\zeta,a,t) + c_{LM} I_{LM}(\zeta,a,t) + c_{PCR} I_{PCR}(\zeta,a,t)\big) da\, d\zeta \tag{6.4}$$

Where $c_D$, $c_T$, $c_{LM}$, and $c_{PCR}$ represent the human-to-mosquito infectiousness for untreated symptomatic infection, treated symptomatic infection, asymptomatic LM-detectable infection and asymptomatic PCR-detectable infection, respectively. Due to the quicker development of *P. vivax* gametocytes compared to *P. falciparum*, there is assumed to be no delay between infection and infectiousness in humans.

*Larval development*

For both *P. falciparum* and *P. vivax* the larval stage model, shown in the following equations, is based on the previously described model in White et al. 2011 (6). Female adult mosquitoes lay eggs at a rate $\beta_L$. Upon hatching from eggs, larvae progress through early and late larvae stages ($E$ and $L$ compartments) before developing into to the pupal stage $P_L$. Adult female mosquitoes emerge from the pupal stage in equation ( 6.1, which is calculated as $\beta = 0.5\,\frac{P_L}{d_P}$.

$$\begin{aligned}
\frac{dE}{dt} &= \beta_L(S_M + E_M + I_M) - \mu_E\left(1 + \frac{E+L}{K}\right)E - \frac{E}{d_E} \\
\frac{dL}{dt} &= \frac{E}{d_E} - \mu_L\left(1 + \gamma\frac{E+L}{K}\right)L - \frac{L}{d_L} \\
\frac{dP_L}{dt} &= \frac{L}{d_L} - \mu_P P_L - \frac{P_L}{d_P}
\end{aligned} \tag{6.5}$$

The duration of each larval stage is represented by $d_E$, $d_L$ and $d_P$. The larval stages are regulated by density-dependent mortality rates, with a time-varying carrying-capacity, $K$, that represents the ability of the environment to sustain breeding sites through different periods of the year and with the density of larvae in relation to the carrying-capacity regulated by a parameter $\gamma$. Since seasonality in transmission dynamics was not modelled at the country level in this analysis, the carrying capacity was assumed to be constant throughout the year. The carrying capacity determines the mosquito density and hence the baseline transmission intensity in the absence of interventions. It is calculated as:

$$K = M_0 \frac{2d_L\mu_0(1 + d_P\mu_P)\gamma(\lambda_M + 1)}{\left(\frac{\lambda_M}{\mu_L d_E} - \frac{1}{\mu_L d_L} - 1\right)} \qquad (6.6)$$

Where $M_0$ is the initial female mosquito density, $\mu_0$ is the baseline mosquito death rate and $\lambda_M$ is defined as:

$$\lambda_M = -0.5\left(\gamma\frac{\mu_L}{\mu_E} - \frac{d_E}{d_L} + (\gamma - 1)\mu_L d_E\right) + \sqrt{0.25\left(\gamma\frac{\mu_L}{\mu_E} - \frac{d_E}{d_L} + (\gamma - 1)\mu_L d_E\right)^2 + \gamma\frac{\beta_L\mu_L d_E}{2\mu_E\mu_0 d_L(1 + d_P\mu_P)}} \qquad (6.7)$$

In this equation, the number of eggs laid per day, $\beta_L$, is defined as:

$$\beta_L = \frac{\beta_{L_{max}}\,\mu e^{-\mu/f_R}}{\mu\left(e^{\mu/f_R} - 1\right)\left(1 - e^{-\mu/f_R}\right)} \qquad (6.8)$$

Where $\beta_{L_{max}}$ is the maximum number of eggs per oviposition per mosquito. The adult mosquito death rate $\mu$ and the mosquito feeding rate $f_R$ are affected by the use of ITNs and further described in the following section on modelling vector control. Full details on the derivation of the egg-laying rate $\beta_L$ and the carrying capacity $K$ have been previously published (6).

#### 7. Modelling the impact of insecticide-treated nets (ITNs)

ITNs are modelled as described previously (1, 4). Mosquito population and transmission dynamics are affected by the use of ITNs in four ways: the mosquito death rate is increased, the feeding or gonotrophic cycle is increased, the proportion of bites taken on protected and unprotected people is changed, and the proportion of bites taken on humans relative to animals is affected. The probability that a blood-seeking mosquito successfully feeds on a human (as opposed to being repelled or killed) will depend on species-dependent bionomics and behaviors of the mosquito, as well as the anti-vectoral interventions present in the human population. Parameter values can be found in **Table S3**.

*Mosquito feeding behavior*

In the model there are 4 possible outcomes of a mosquito feeding attempt:

1. The mosquito bites a non-human host
2. The mosquito attempts to bite a human host but is killed by the ITN before biting
3. The mosquito successfully feeds on a human host and survives that feeding attempt
4. The mosquito attempts to bite a human host but is repelled by the ITN without feeding, and repeats the attempt find a blood meal source.

We define the probability of a mosquito biting a human host during a single attempt as $y_i$, the probability that a mosquito bites a human host and survives the feeding attempt as $w_i$, and the probability of a mosquito being repelled without feeding as $z_i$. These probabilities exclude natural vector mortality, so that for a population without protection from ITNs (e.g. prior to their introduction), $y_1 = w_1 = 1$ and $z_1 = 0$.

The presence of ITNs modifies these probabilities of surviving a feeding attempt or being repelled without feeding. Upon entering a house with ITNs, mosquitoes can experience three different outcomes: being repelled by the ITN without feeding (probability $r_N$), being killed by the ITN before biting (probability $d_N$), or feeding successfully (probability $s_N$). It is assumed that all biting attempts inside a house occur on humans. The repellency of ITNs in terms of the insecticide and barrier effect decays over time, giving the following probabilities:

$$r_N = (r_{N0} - r_{NM}) \exp(-t\gamma_N) + r_{NM} \quad (7.1)$$

$$d_N = d_{N0} \exp(-t\gamma_N) \quad (7.2)$$

$$s_N = 1 - r_N - d_N \quad (7.3)$$

Where $r_{N0}$ is the maximum probability of a mosquito being repelled by a bednet and $r_{NM}$ is the minimum probability of being repelled by a bednet that no longer has insecticidal activity and possibly holes reducing the barrier effect. $\gamma_N$ represents the rate of decay of the effect of ITNs over time $t$ since their distribution and is calculated as $\frac{\log(2)}{LLIN\ half\text{-}life}$. The killing effect of ITNs decreases at the same constant rate from a maximum probability of $d_{N0}$. In model simulations, ITNs are distributed every three years.

With $i = 1$ representing the population not covered by an ITN and $i = 2$ representing the population covered by an ITN, this gives the following probabilities of successfully feeding, $W$, and being repelled without feeding, $Z$, during a single feeding attempt on a human:

$$W = \sum_{i=1}^{2} w_i c_i \qquad w = \begin{cases} 1 & if\ i = 1 \\ 1 - \Phi_b + \Phi_b s_N & if\ i = 2 \end{cases} \quad (7.4)$$

$$Z = \sum_{i=1}^{2} z_i c_i \qquad z = \begin{cases} 0 & if\ i = 1 \\ \Phi_b r_N & if\ i = 2 \end{cases} \quad (7.5)$$

Where $c_i$ is the proportion of the population in the respective group, and $\Phi_b$ is the proportion of bites taken on humans in bed, which was derived in previous publications (4).

During a single feeding attempt (which may be on animals or humans), the average probabilities of mosquitoes feeding or being repelled without feeding, $\bar{W}$ and $\bar{Z}$, are then:

$$\bar{W} = 1 - Q_0 + Q_0 W \quad (7.6)$$

$$\bar{Z} = Q_0 Z \quad (7.7)$$

Where $Q_0$ is the proportion of bites taken on humans in the absence of any vector control intervention.

*Effect of ITNs on mosquito mortality*

The average probability of mosquitoes being repelled without feeding in the model affect the mosquito feeding rate, $f_R$, as follows:

$$f_R = \frac{1}{\frac{\delta_1}{(1-\bar{Z})} + \delta_2} \tag{7.8}$$

Where $\delta_1$ is the time spent looking for a blood meal in the absence of vector control, and $\delta_2$ is the time spent resting between blood meals, which is assumed to be unaffected by ITN usage.

The average probabilities of feeding or being repelled also affect the probability of surviving the period of feeding, $p_1$, as follows:

$$p_1 = \frac{\bar{W}\exp(-\mu_0\delta_1)}{1-\bar{Z}\exp(-\mu_0\delta_1)} \tag{7.9}$$

Where $\mu_0$ is the baseline mosquito death rate in the absence of interventions.

The probability of surviving the period of resting, $p_2$, is not affected by ITNs:

$$p_2 = \exp(-\mu_0\delta_2) \tag{7.10}$$

This allows to calculate the mosquito mortality rate affecting mosquito population dynamics in the set of equations ( 6.1:

$$\mu = -f_R \ln(p1 * p2) \tag{7.11}$$

*Effect of ITNs on the force of infection acting on humans and mosquitoes*

In the presence of ITNs, the anthropophagy (the proportion of successful bites which are on humans) of mosquitoes is represented by parameter $Q$. This is affected by ITN usage as follows:

$$Q = 1 - \frac{1-Q_0}{\bar{W}} \tag{7.12}$$

Further details on the assumptions in this calculation can be found in an earlier publication (4).

This then gives the biting rate on humans, $\alpha$, as shown in the equations for the force of infection experienced by humans (equations ( 4.2-( 4.4) and by mosquitoes (equations ( 6.2 and ( 6.4):

$$\alpha = Qf_R\frac{w_{int}}{W} \tag{7.13}$$

*Effect of ITNs on larval development*

The mosquito death rate $\mu$ and the feeding rate $f_R$ also influence the calculation of the carrying capacity $K$ and the egg-laying rate $\beta_L$ in equations ( 6.6 and ( 6.8, thereby affecting larval development.

**Table S3**. Mosquito model and ITN parameter. Full details on parameter values can be found in the original publications (1, 2, 4, 6), including references and intervals for the prior and posterior distributions for fitted parameters (median values of the posterior distribution are used in model simulations).

| | | ***P. falciparum* (*Anopheles gambiae s.s.*)** | ***P. vivax* (*Anopheles punctulatus*)** |
|---|---|---|---|
| **Infectiousness of humans to mosquitoes** | | | |
| Lag from parasites to infectious gametocytes | $\tau_1$ | 12.5 days | - |
| Untreated clinical disease | $c_D$ | 0.068 | 0.8 |
| Treated clinical disease | $c_T$ | 0.022 | 0.4 |
| Sub-patent infection | $c_U$ | 0.0062 | - |
| Parameter for infectiousness of asymptomatic infection | $\gamma_1$ | 1.82425 | - |
| Light microscopy-detectable infection | $c_{LM}$ | - | 0.1 |
| PCR-detectable infection | $c_{PCR}$ | - | 0.035 |
| **Mosquito Population Model** | | | |
| Daily mortality of adult mosquitoes with no interventions | $\mu_0$ | 0.132 | 0.167 |
| Extrinsic incubation period | $\tau_M$ | 10 days | 8.4 days |
| **Larval model** | | | |
| Early instar larval developmental period | $d_E$ | 6.64 days | 6.64 days |
| Late instar developmental period | $d_L$ | 3.72 days | 3.72 days |
| Pupal developmental period | $d_P$ | 0.643 days | 0.643 days |
| Daily mortality rate of early-stage larvae (density dependent) | $\mu_E$ | 0.0338 | 0.0338 |
| Daily mortality rate of late-stage larvae (density dependent) | $\mu_L$ | 0.0348 | 0.0348 |
| Daily mortality rate of pupae (density independent) | $\mu_P$ | 0.249 | 0.249 |
| Effect of density dependence on late instars relative to early instars | $\gamma$ | 13.25 | 13.25 |
| Maximum number of eggs per oviposition per mosquito | $\beta_{L_{max}}$ | 21.2 | 21.2 |
| **Mosquito behavior** | | | |
| Mean duration of host-seeking in the absence of vector control interventions | $\delta_1$ | 0.69 days | 0.69 days |
| Mean duration of resting between blood meals | $\delta_2$ | 2.31 days | 2.31 days |

| | | | |
|---|---|---|---|
| Proportion of bites taken on humans (anthropophagy) in the absence of vector control interventions | $Q_0$ | 0.92 | 0.5 |
| Proportion of bites taken on humans indoors and in bed | $\Phi_b$ | 0.89 | 0.9 |
| **Effect of insecticide-treated nets (ITNs)** | | | |
| Maximum probability of a mosquito being repelled by a ITN with full insecticidal and barrier effect | $r_{N0}$ | 0.56 | 0.6 |
| Minimum probability of a mosquito being repelled by a ITN after decay | $r_{NM}$ | 0.24 | 0.2 |
| ITN half-life | - | 2.64 years | 2.64 years |
| Maximum probability of a mosquito being killed by a ITN with full insecticidal and barrier effect | $d_{N0}$ | 0.41 | 0.3 |

#### 8. Assumptions in model outcomes

*Model dynamics over time*

To represent long-term reductions in clinical burden, model simulations were run until a new equilibrium was reached post-intervention for all ITN usage levels, which corresponded to 75 years for *P. falciparum* and 175 years for *P. vivax*. As shown in **Figure S2**, when ITNs are continuously distributed over time, clinical incidence outcomes initially fluctuate before reaching a long-term equilibrium due to various effects on population immunity and mosquito population dynamics in the model. For example, in high-transmission *P. falciparum* settings, clinical incidence experiences a steep initial decline after ITN introduction, before gradually rebounding to an equilibrium value (**Figure S2A**). In the *P. vivax* model, stabilization at an equilibrium transmission level was further delayed due to the presence of hypnozoites in a deterministic framework, whereby even an extremely small reservoir could lead to rebounds in clinical infections after decades. To limit *P. vivax* simulations to a computationally feasible time period, we prevented this rebound by introducing the assumption that once a hypnozoite prevalence of less than 1 in 100,000 is reached in the population, the reservoir is further depleted and cannot lead to a renewed chain of transmission (**Figure S2B**).

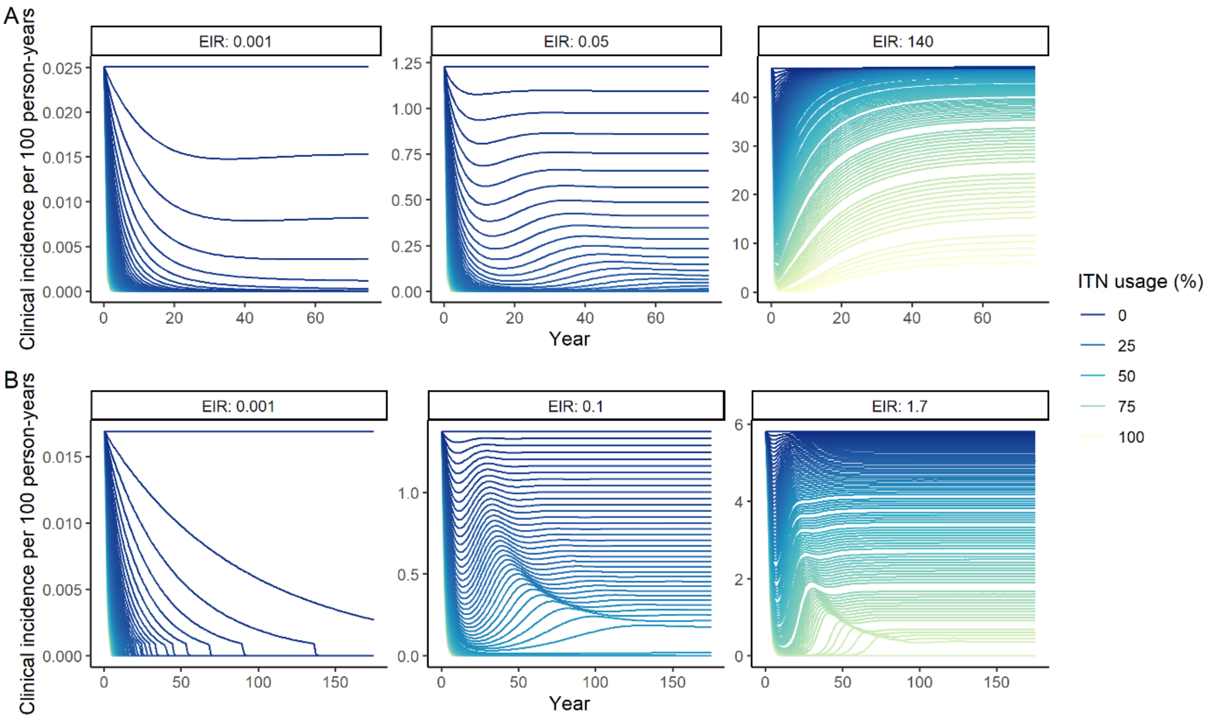


**Figure S2**. Modelled impact of ITN usage on clinical incidence over time for three representative entomological inoculation rates (EIR) for A) *P. falciparum* and B) *P. vivax*. EIRs represent the minimum, median, and maximum EIRs of the global population distribution. Lines represent increments of 1% of ITN usage.

*Clinical incidence and assumptions about case detection*

We simulated clinical incidence assuming cases would be detected through weekly active case detection (ACD). ACD represents a more sensitive method to assess disease burden and was used in the majority of studies used to calibrate *P. falciparum* and *P. vivax* models (2, 4). This assumption results in higher case incidence than reported case numbers because not everyone seeks care at a health clinic for a clinical episode (4). As estimated in previous publications, weekly ACD was assumed to detect 72.3% and 13.4% of all *P. falciparum* and *P. vivax* clinical cases detected by daily ACD, respectively (2, 4, 7).

#### Country-level data and modelling assumptions on the global malaria distribution

To represent the global distribution of malaria, a *P. falciparum* prevalence in 2-10 year-olds (*Pf*$PR_{2\text{-}10}$) (2000) raster layer (8)was clipped to a *P. falciparum* transmission spatial limits (2010) raster layer (9) obtained from the Malaria Atlas Project. Country shapefiles, obtained from geoBoundaries (10), were overlaid on prevalence estimates, and the mean *Pf*$PR_{2\text{-}10}$ within each boundary was calculated. A similar process was completed for *P. vivax* using *Pv*$PR_{0\text{-}99}$ (2000) and *P. vivax* transmission spatial limits (2010) raster layers (11). WorldPop gridded 2000 global population estimates (12) were summed within boundaries to output the total population at risk of malaria infection living within each country. For both species, parasite prevalence was then matched to modelled entomological inoculation rates (EIR) associated with the closest prevalence estimate. The group of countries with lowest transmission intensity included those with an EIR of 0.001 or lower.

In our analysis, we assumed that most of sub-Saharan Africa was not endemic for *P. vivax*, because *P. vivax* prevalence and incidence could not be estimated (11). Even though there is evidence for low-level *P. vivax* endemicity throughout the continent, there is no routine surveillance for non-*P. falciparum* cases and the prevalence of the Duffy-negative phenotype among African populations is protective against endemic transmission of *P. vivax* (11). Therefore, our estimates for the population at risk of *P. vivax* malaria do not include much of sub-Saharan Africa (except the Horn of Africa).

Although model simulations were matched to country-level prevalence, we did not aim to capture the wide geographic variation in malaria epidemiology in detail. For example, in all simulations with the *P. vivax* model, we fixed the relapse and hypnozoite clearance rates, based on the original parameter values used in the calibrated model in Papua New Guinea (2). The timings of relapse are thought to follow different patterns in different geographical areas, with a particular distinction between tropical strains relapsing quickly after initial infection and temperate strains relapsing only after 6-12 months (13). Nevertheless, projections from the model calibrated to sub-national Papua New Guinean data were also shown to be in line with global epidemiological patterns at various prevalence levels (2). Similarly, we did not account for the geographic variation in dominant malaria vector species, which are particularly diverse across *P. vivax* endemic areas (14).

In all model simulations and analyses, we assumed infections with the two parasite species to be independent, in line with the presentation of estimates from Malaria Atlas Project. Therefore, in each setting we considered total malaria cases to represent the sum of modelled *P. falciparum* and *P. vivax* cases, total malaria prevalence to represent the sum of *P. falciparum* and *P. vivax* parasite prevalence, and the total EIR to represent the sum of average *P. falciparum*- and *P. vivax*-infectious bites per person year. With the geographical areas endemic for the two species overlapping in many locations, we assumed the population at risk of malaria in each setting to represent the higher of the population at risk of *P. falciparum* or of *P. vivax*.

#### Relationship between distribution and usage of insecticide-treated nets (ITNs)

As described in the manuscript, the non-linear relationship between costs and ITN usage was accounted for by converting the modelled population usage into the required number of ITNs to be distributed to achieve this usage. For this, a published methodology was used; full assumptions and definitions can be found in the original publication (15). Equations are detailed below and parameter values for the application in this paper are summarized in **Table S4**.

Firstly, the simulated ITN usage was converted into ITN population access based on observed ITN use rates. By definition:

$$ITN\ access = \frac{ITN\ usage}{ITN\ use\ rate}$$

Since access in the population cannot exceed 1, the modelled ITN usage could not be higher than the assumed use rate.

Secondly, a Loess curve was fitted to 2020 data on net access and nets per capita per country-month from across Africa, reproducing a similar relationship as shown in the original publication (**Figure S3**) (15). The net access derived for a given usage was then converted into nets per capita using the Loess curve. We extrapolated the trend for higher access levels and assumed that all access levels below the minimum observed would require the same nets per capita (i.e. the same cost) to achieve.

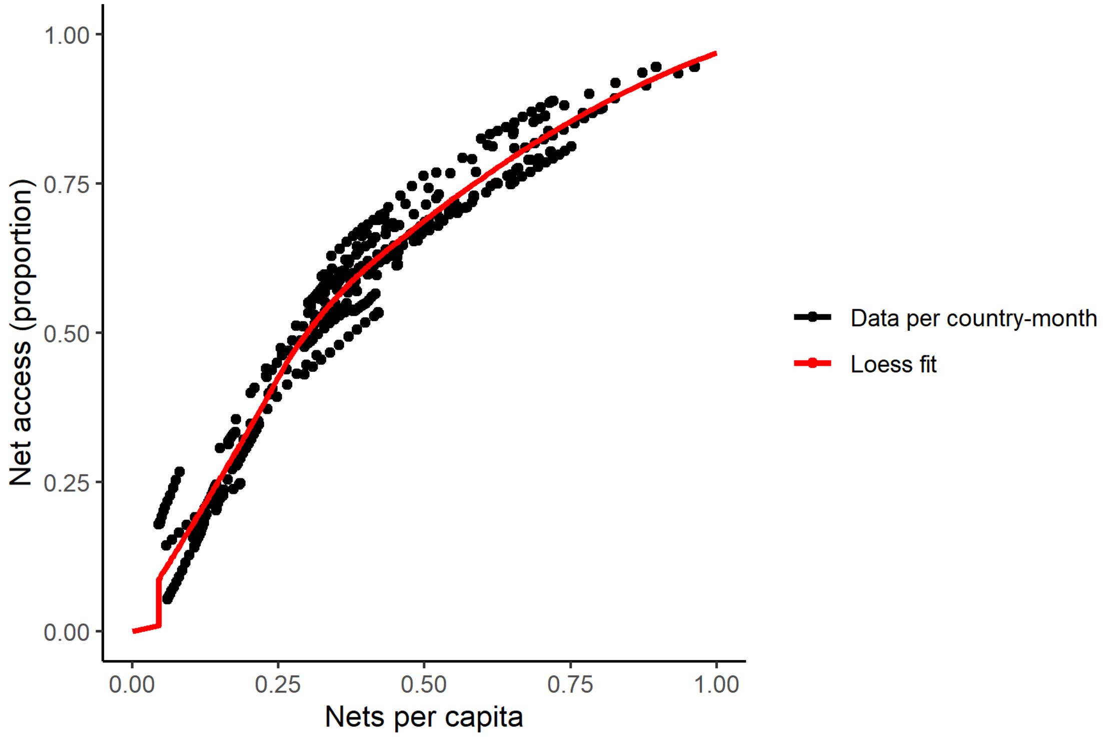


**Figure S3**. Relationship between access and nets per capita in 2020 (generated from data in Bertozzi-Villa *et al.*, 2021).

Lastly, the nets per capita were converted into the nets distributed per person-year, accounting for net retention over time and assuming a distribution frequency of every 3 years. Like in the original publication, ITNs were assumed to be lost from the population following a smooth compact function after distribution, so that the proportion of nets retained over time, $p(t)$, equals:

$$p(t) = \begin{cases} e^{\kappa - \kappa / 1-\left(\frac{t}{\tau}\right)^2} & if\ t < \tau \\ 0 & if\ t \geq \tau \end{cases}$$

Where $\kappa$ is a fitted rate parameter estimated from the data in the original publication. $\tau$ determines the time by which no nets are retained in the population, and was estimated from the assumed net half-life, as follows:

$$\tau = \frac{ITN\ half - life}{\sqrt{1 - \frac{\kappa}{\kappa - \ln(0.5)}}}$$

Integrating the net loss function over a distribution cycle then allows to derive the annual nets distributed per capita:

$$Nets\ distributed\ per\ capita\ per\ year = \frac{nets\ per\ capita}{DF * \int_0^{DF} p(t)\ dt}$$

Where $DF$ represents the distribution frequency.

**Table S4**. Parameter values for the insecticide-treated net (ITN) costing conversion.

| PARAMETER | SYMBOL | VALUE | SOURCE |
|---|---|---|---|
| **ITN usage** | - | Varies in simulations | - |
| **ITN use rate (proportion)** | - | 0.84 | Median across African countries in 2019 (15) |
| **ITN half-life (years)** | - | 1.64 | Median across African countries in 2020 (15) |
| **ITN distribution frequency (years)** | $DF$ | 3 | World Malaria Report (16) |
| **Net loss function rate parameter** | $\kappa$ | 20 | Bertozzi-Villa *et al.*, 2021 (15) |

#### Optimization model

The mathematical problem consists in finding the allocation *b* of ITNs that minimizes global malaria cases, i.e. the sum of the product between the population $p_i$ times the clinical incidence $cinc_i$ for each EIR setting *i*. In the objective function, we also allow for the option of placing a positive contribution on settings reaching a pre-elimination phase (defined as a clinical incidence of less than 1 case per 1000 persons at risk) in addition to minimizing the global malaria case burden. This premium accounts for potential benefits of reaching low levels of malaria transmission that go beyond the reduction in cases, e.g. general health system strengthening. For each setting reaching pre-elimination, the total remaining cases are reduced by a proportion *w* of the total cases averted by the ITN allocation (compared to total cases at baseline/without interventions), *C*. *w* therefore represents the weighting placed on pre-elimination in a setting relative to total case reduction. In the scenario optimized for case reduction, this weight equals 0.

This optimization must respect the budget constraint that the cost of ITNs distributed at each EIR setting $b_i$ must be less than or equal to the total budget *B*, with *c* being the cost of a single pyrethroid-treated net. In addition, the ITN usage $b_i^*$ in each setting *i* must be between 0% and an upper limit of 80%, which is a common target for universal access (17). Notice that in our model, ITN distributed $b_i$ is not the same as ITN usage $b_i^\star$, because only a fraction of ITNs distributed will be used over time. We represent with $f(b_i)$ the function that maps ITNs distributed into ITNs used (see "Relationship between distribution and usage of insecticide-treated nets" for more details on this function):

$$\min_{b \in \mathbb{R}^n} \left[ \sum_{i}^{n} cinc_i * p_i \; - \; w * C * \sum_{i=1}^{n} j_i \right]$$

$$\text{s.t.} \qquad \sum_{i=1}^{n} b_i * c \le B$$

$$0 \le b_i^\star \le 0.8 \;\; \forall \, i = 1, \dots, n$$

$$C = Cases\ at\ baseline - \sum_{i}^{n} cinc_i * p_i$$

$$j_i = \begin{Bmatrix} 1, & cinc_i < 1/1000 \\ 0, & cinc_i \ge 1/1000 \end{Bmatrix}$$

$$b_i^\star = f(b_i)$$

$$\text{for all } i = 1, \dots, n$$

Optimization was performed using generalized simulated annealing using the *GenSA* R package (v.1.1.7.) (18). *GenSA* can receive a non-linear objective function and searches an inputted search space for the global minimum. The function can tolerate a field which contains multiple local minima by simulating an annealing process using the stochasticity of a temperature parameter to escape local minima and continue the search for a global minimum (18). Because many different combinations of ITN usage levels across different settings can lead to small case numbers, our objective function has many local minima. Therefore, we decided, as suggested in (18), to use a high value of $10^6$ for the temperature and to increase the maximum number of iterations from the default values of $5 * 10^4$ to $5 * 10^6$.

Since this version of the algorithm is not designed for constrained optimization, we transformed the problem into an unconstrained optimization by introducing a penalty term in the objective function. The unconstrained problem without the pre-elimination premium can be represented as:

$$\min_{b \in \mathbb{R}^n} \sum_{i=1}^{n} cinc_i \cdot p_i + F(b)$$

$$\text{s.t.} \qquad 0 \le b_i^\star \le 0.8 \;\; \forall\, i = 1, \dots, n$$

$$\text{with} \quad F(b) = \begin{cases} 0 \quad if \sum_i b_i * c \le B \\ 10^{10} \, if \sum_i b_i * c > B \end{cases}$$

$$\text{and} \qquad b_i^\star = f(b_i)$$

Namely, the objective function will assume a very high value in all cases where the budget constraint is not respected. In this way, the simulated annealing algorithm would discard all solutions outside of the budgetary constraints.

The search space was built using the *akima* package (v.0.6-2.2, Akima and Gebhardt 2021), to construct two 3D surfaces of clinical incidence model outputs for every combination of bed net usage and EIR (**Figure S4**). The dimensions of the resulting surfaces were 9000 x 9000 points.

The optimization function was run through a range of $B$ from no intervention (starting point) to full coverage, with results indicating the resource allocation combination which most reduced clinical incidence from baseline at each level of funding.

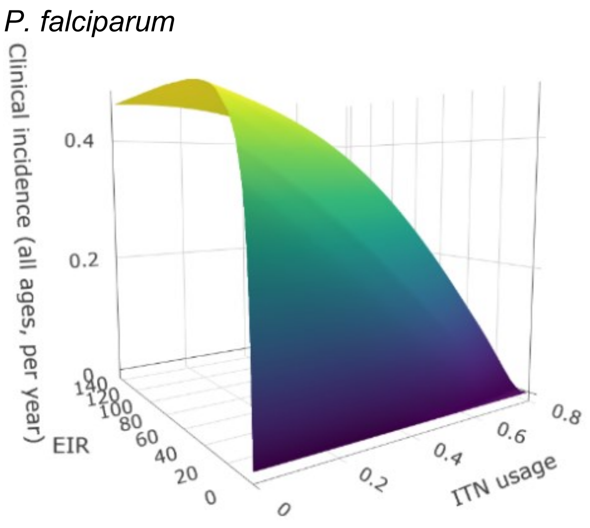


**Figure S4**. Surface plots of EIR vs. ITN usage vs. clinical incidence. Plots were fit using bivariate linear interpolation of gridded data EIR and ITN usage values taken from mathematical model simulations.

### SUPPLEMENTARY FIGURES

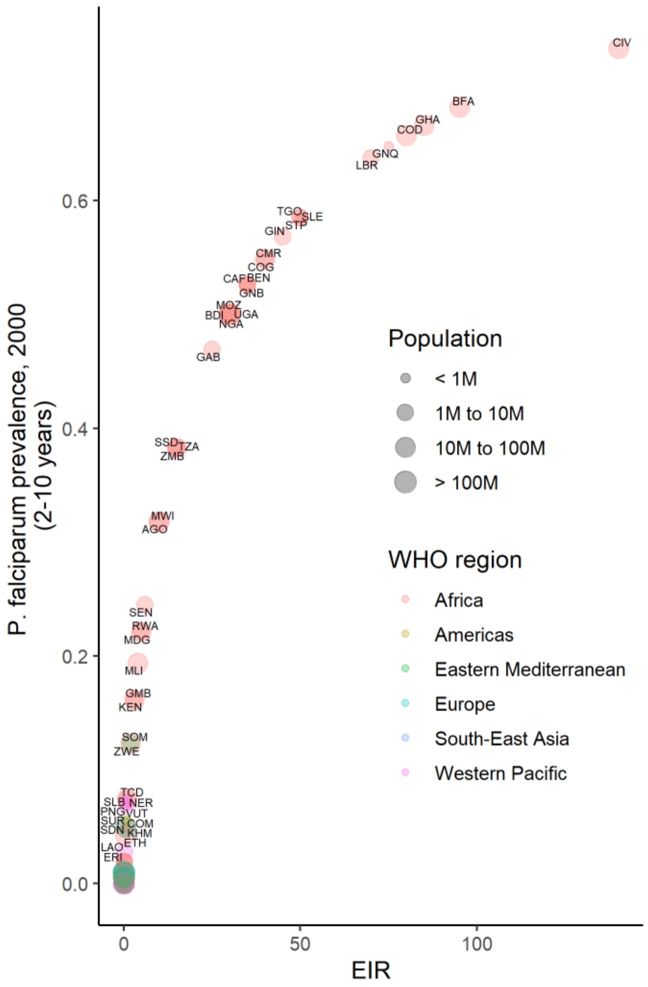


| EIR | n | population | % of global |
|---|---|---|---|
| 0.001 | 22 | 1,435,092,326 | 34.60 |
| 0.010 | 14 | 295,534,501 | 7.13 |
| 0.050 | 5 | 345,639,624 | 8.33 |
| 0.100 | 8 | 1,460,442,975 | 35.21 |
| 0.200 | 3 | 7,411,614 | 0.18 |
| 0.300 | 1 | 5,082,884 | 0.12 |
| 0.500 | 1 | 61,335,932 | 1.48 |
| 0.600 | 3 | 37,273,416 | 0.90 |
| 0.700 | 1 | 459,762 | 0.01 |
| 0.900 | 2 | 5,150,866 | 0.12 |
| 1.000 | 3 | 18,728,450 | 0.45 |
| 2.000 | 2 | 17,737,454 | 0.43 |
| 3.000 | 2 | 30,420,052 | 0.73 |
| 4.000 | 1 | 10,426,400 | 0.25 |
| 5.000 | 2 | 22,962,444 | 0.55 |
| 6.000 | 1 | 8,826,698 | 0.21 |
| 10.000 | 2 | 30,663,795 | 0.74 |
| 15.000 | 3 | 48,254,642 | 1.16 |
| 25.000 | 1 | 1,276,950 | 0.03 |
| 30.000 | 4 | 162,288,085 | 3.91 |
| 35.000 | 3 | 10,821,418 | 0.26 |
| 40.000 | 2 | 19,533,204 | 0.47 |
| 45.000 | 1 | 7,562,499 | 0.18 |
| 50.000 | 3 | 9,014,672 | 0.22 |
| 70.000 | 1 | 2,733,044 | 0.07 |
| 75.000 | 1 | 968,746 | 0.02 |
| 80.000 | 1 | 45,833,588 | 1.11 |
| 85.000 | 1 | 18,554,042 | 0.45 |
| 95.000 | 1 | 11,586,619 | 0.28 |
| 140.000 | 1 | 15,902,142 | 0.38 |

**Figure S5**. Prevalence of *P. falciparum* in children 2-10 years (2000), matched to EIR values by country. Points are sized by total population and colored by World Health Organization region.

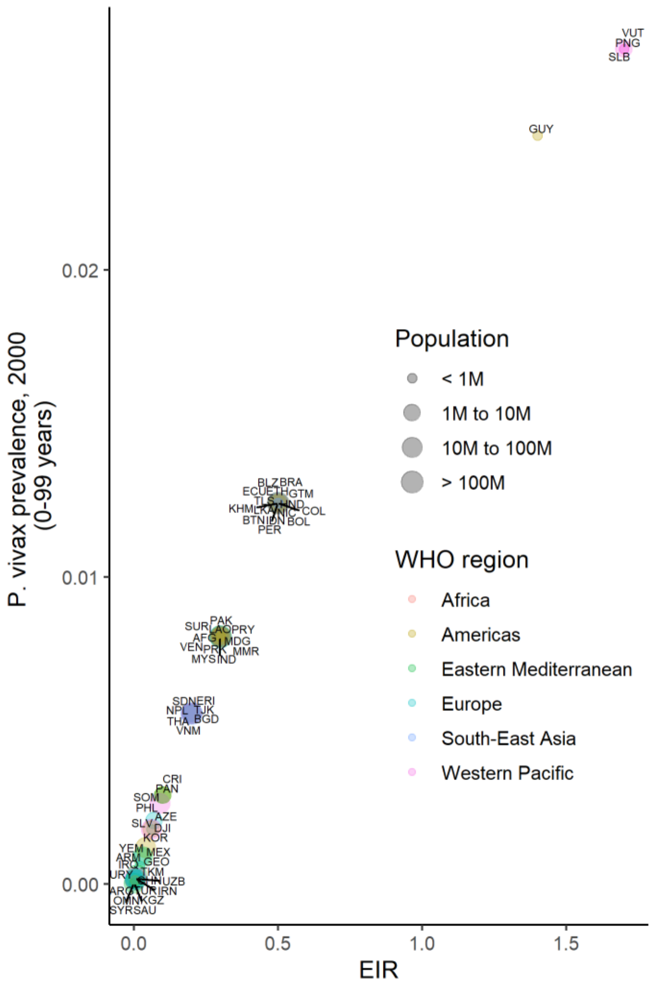


| EIR | n | population | % of global |
|---|---|---|---|
| 0.001 | 3 | 19,930,651 | 0.50 |
| 0.004 | 4 | 109,300,522 | 2.76 |
| 0.006 | 3 | 1,335,451,364 | 33.77 |
| 0.010 | 2 | 24,250,242 | 0.61 |
| 0.020 | 1 | 16,414 | 0.00 |
| 0.030 | 2 | 21,891,874 | 0.55 |
| 0.040 | 1 | 95,187,544 | 2.41 |
| 0.060 | 3 | 49,291,598 | 1.25 |
| 0.070 | 1 | 7,838,269 | 0.20 |
| 0.090 | 1 | 73,308,400 | 1.85 |
| 0.100 | 3 | 12,803,124 | 0.32 |
| 0.200 | 7 | 314,893,676 | 7.96 |
| 0.300 | 11 | 1,322,338,591 | 33.44 |
| 0.500 | 15 | 561,789,177 | 14.21 |
| 1.400 | 1 | 704,403 | 0.02 |
| 1.700 | 3 | 5,395,726 | 0.14 |

**Figure S6**. Prevalence of *P. vivax* in people 0-99 years (2000), matched to EIR values by country. Points are sized by total population and colored by World Health Organization region.

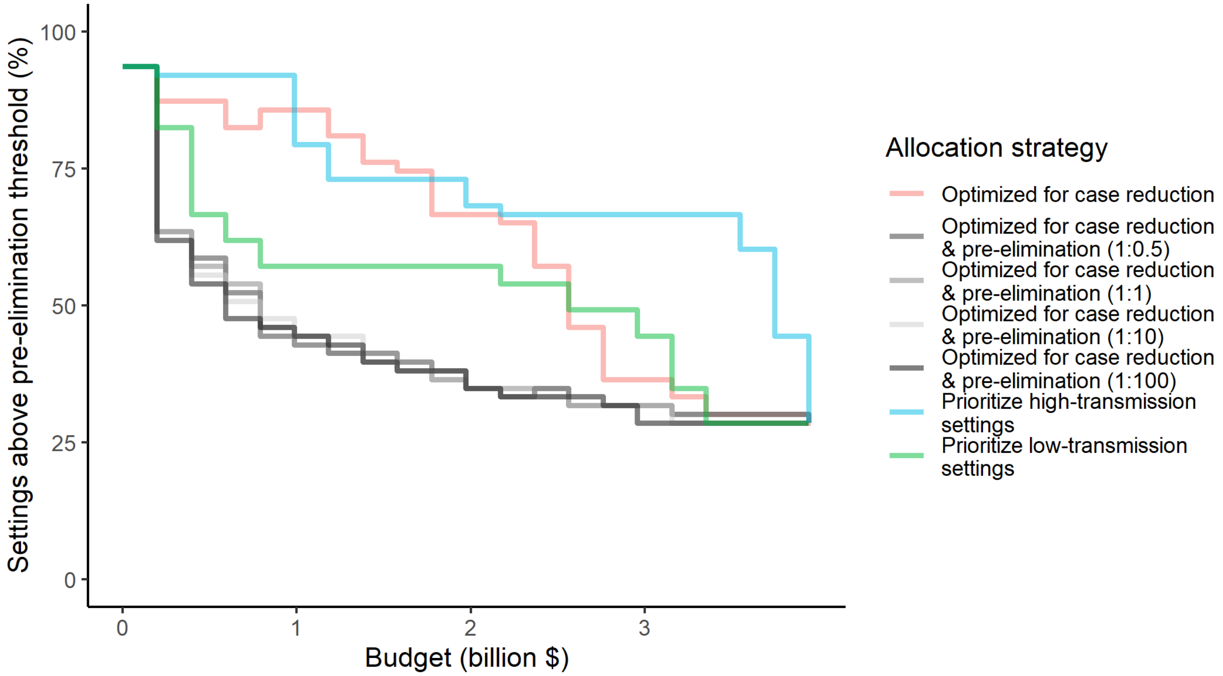


**Figure S7.** Percentage of settings not having reached pre-elimination (<1 case per 1000 population at risk) under different allocation strategies at varying budgets. Budget levels range from 0, representing no usage of insecticide-treated nets, to the budget required to achieve the maximum possible impact. For the strategies optimizing for case reduction and pre-elimination, brackets show the weight placed on averting total cases *vs* on reaching pre-elimination in the optimization (i.e. 1:1 represents equal weight on both).

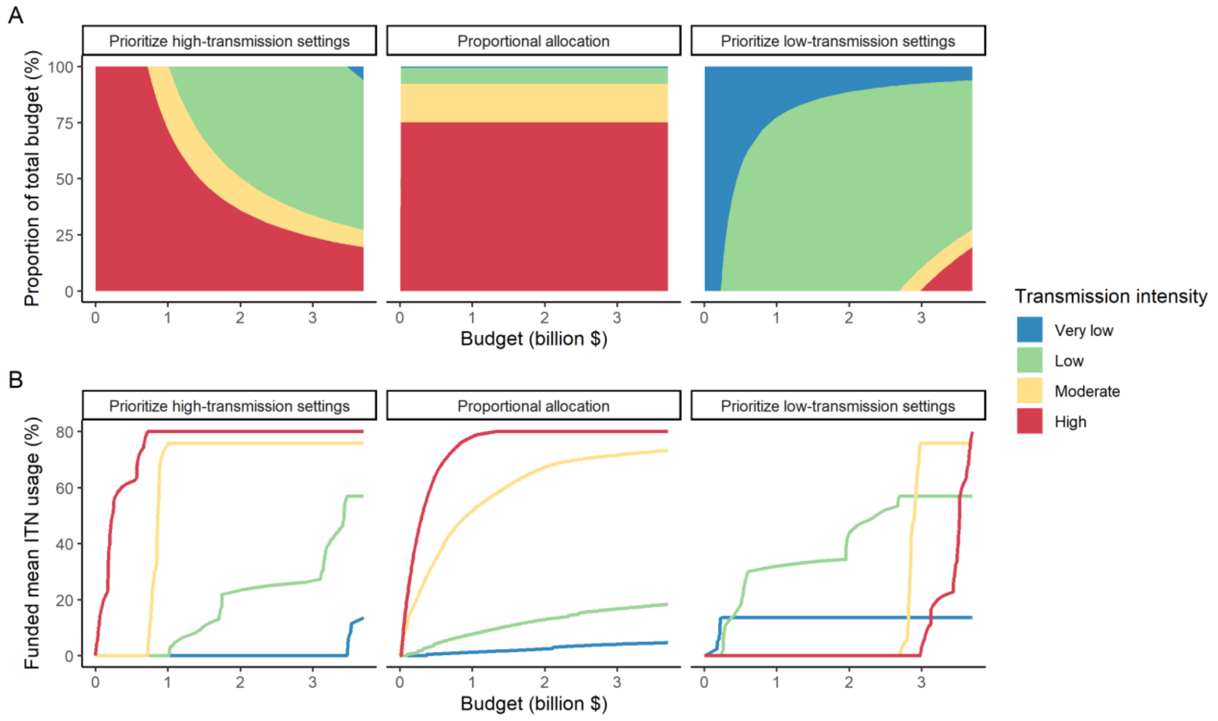


**Figure S8**. Illustration of resource allocation patterns under the three modelled policy strategies at varying budgets. A) percentage of global funding allocated to each transmission setting. B) mean funded ITN usage in each transmission setting. Transmission intensity groups represent transmission settings with varying population sizes proportional to the global distribution.

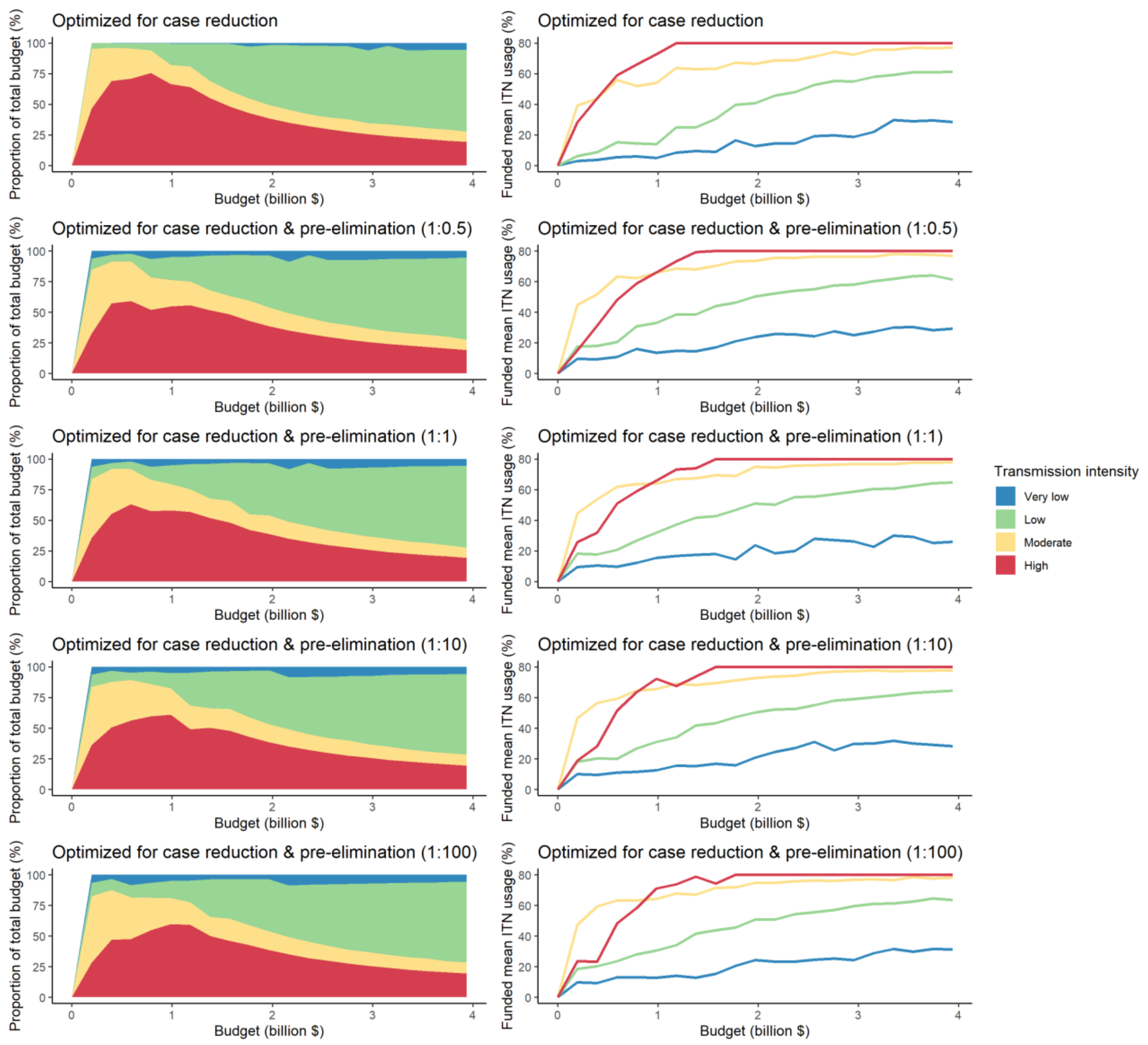


**Figure S9.** Optimal strategies for funding allocation across settings to minimize malaria case burden (top panels) and to minimize malaria cases and increase the number of settings reaching a pre-elimination phase at varying budgets. Panels show the proportion of the budget allocated and the resulting mean population usage of insecticide-treated nets (ITNs) across settings of different transmission intensity. For the strategies optimizing for case reduction and pre-elimination, brackets show the weight placed on averting total cases *vs* on reaching pre-elimination in the optimization (i.e. 1:1 represents equal weight on both).

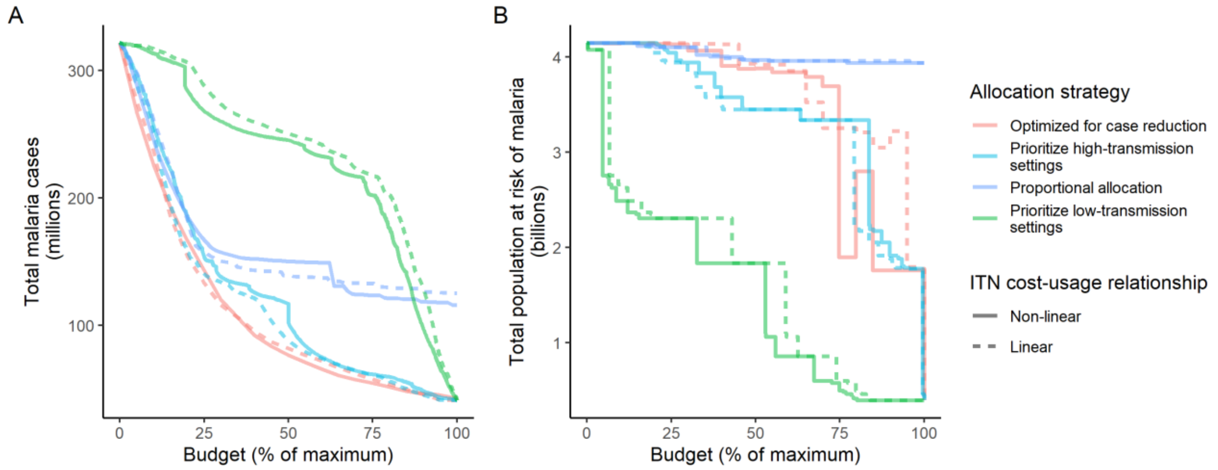


**Figure S10: Influence of different assumptions about the relationship between the cost and population usage of insecticide-treated nets (ITNs) on the impact of the allocation strategies.** The global clinical malaria cases (panel A) and the population at risk of malaria (panel B) under different allocation strategies are shown at varying budgets. Results with the more realistic non-linear assumption are presented throughout the main manuscript. Budget levels are expressed relative to the maximum budget required to achieve the largest possible impact with ITNs, but note that this maximum budget was different depending on the ITN costing assumption ($3,698,727,241 for non-linear *vs.* $6,902,923,309 for linear).

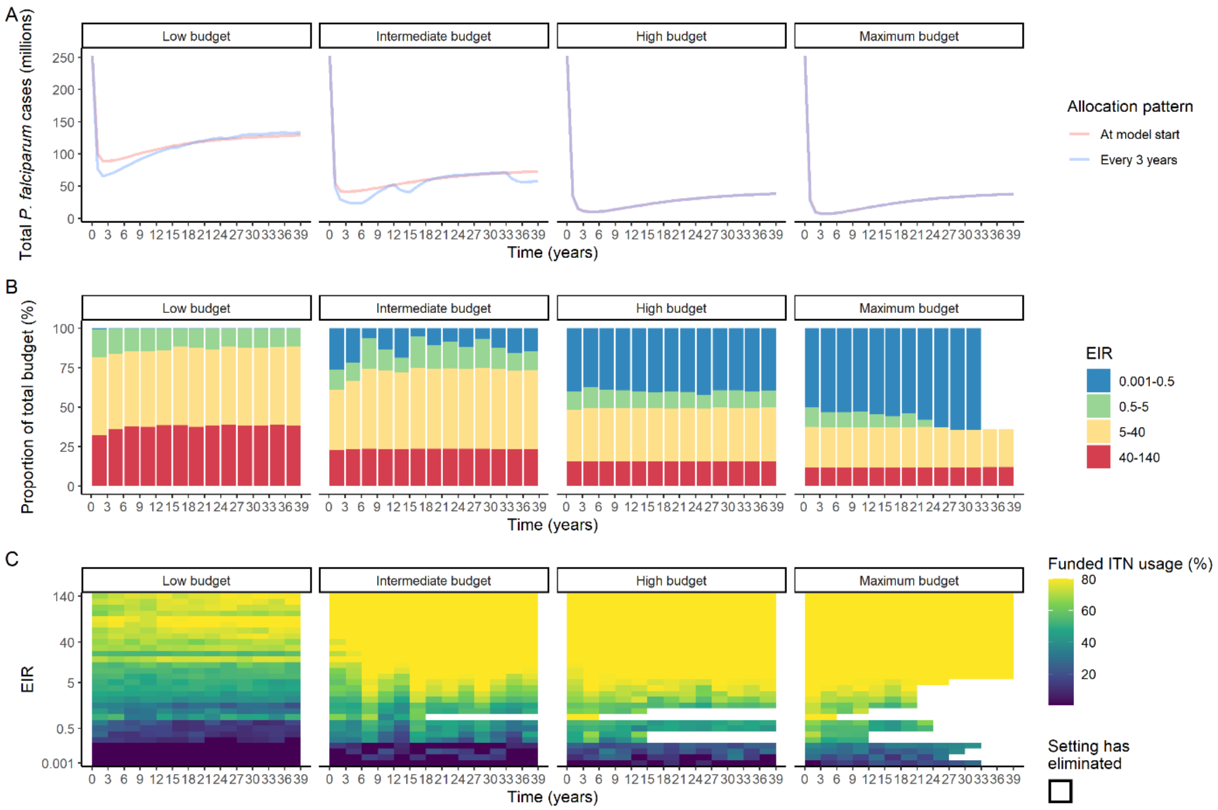


**Figure S11**. Resource allocation patterns over time for *P. falciparum*. Panel A shows the number of cases over time for re-allocation of insecticide-treated nets (ITNs) every 3 years compared to one-time allocation of a constant ITN usage to minimize the final (year 39) case burden. Panels B and C show the optimal allocation pattern for each 3-year distribution cycle across settings of different transmission intensity. The maximum budget was 26.6 million for ITN distributions every 3 years over 39 years.

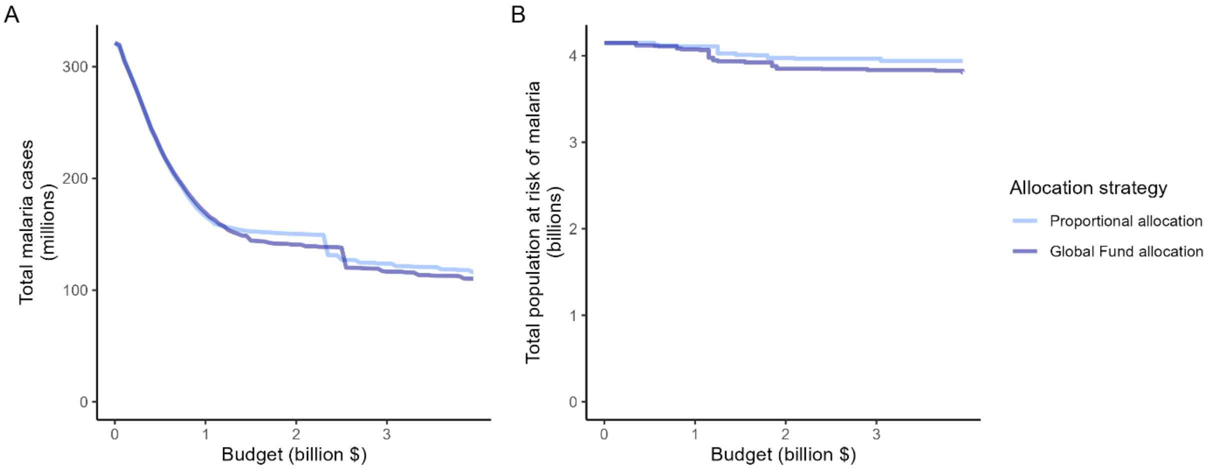


**Figure S12.** Impact of the proportional allocation strategy and the 2020-2022 Global Fund allocation on global malaria cases (panel A) and the total population at risk of malaria (panel B) at varying budgets. Both strategies use the same algorithm for budget share allocation based on malaria disease burden in 2000-2004, but the Global Fund allocation additionally involves an economic capacity component and specific strategic priorities (19).
